# Effect of water intake and storage time on protein concentration and enzyme AChE activity in erythrocyte and plasma blood samples of healthy individuals

**DOI:** 10.1101/2020.10.02.20205823

**Authors:** Snežana Jovičić

## Abstract

**Background:** Storage time influence concentration levels of blood biomarker. This study aimed to assess the effect of **water intake prior sampling and storage time** on protein concentration, enzyme AChE activity, inhibitor efficacy and to build an efficient inhibitor calibration curve in healthy individuals.

**Methods:** Data analysis was performed on 11 participants. Study utilizes substrate acetylcholine chloride and inhibitors **BW284c51 (0.01mM) and GUK-987 (0.1mM)**. Calibration curve ranging from 10^-1^ to 10^-38^ mM was build for inhibitor GUK-987 and GDK-510.

Data analysis is carried out with **Microsoft Excel 2007**. Data analysis was performed via **IBM SPSS Statistical Software v23.0**. Descriptive statistics and parametric statistical tests were assessed for 0, 90, 91 and 92 days of storage in Plasma and Erythrocyte samples. Workflow of building calibration curve and the most efficient inhibition concentration is assessed.

**Results:** **Water intake and storage time** have effect on in vitro protein concentration, activation/inhibition of enzyme AChE activity in Plasma and Erythrocyte samples. However, 100% inhibitor efficacy is maintained for inhibitor GUK-987 in Plasma samples and inhibitor BW284c51 in Erythrocyte samples. The most efficient inhibitor concentration is determined.

**Conclution:** Significant changes and variable association have been estimated between protein concentration, activation/inhibition of enzyme AChE activity, as a cause of water intake and storage time. Taking all these factors into account for further research is important for disease prevention and human wellbeing.

## 1. Introduction

Laboratory equipment, accommodation, organization of work and labor protection measures are of key importance in scientific and clinical research [1]. Differences in working task employ efficient working organization for easy and fast communication between employees. Efficient utilization of work place, quality and complete safety equipment, professional staff training are the basis of good and quality measures on workplace. Safety measures, laboratory type, equipment, chemicals, methodology of work, instrument calibration represent the basis for conducting research on specific biological samples [2]. Quality and efficacy of the results are dependent on good organization skill and keeping administrative records of samples and procedures [3].

Usually, human biological material is taken from diverse admission/clinical departments and transported for laboratory analysis in clinical/scientific settings [3]. These materials needs to be properly marked, sorted, storage and transported in order to obtain precise and proper conclusions. Basic issued rules are implemented in practice [2]. This set of rules can have adverse effect on analyzed analite concentration and specificity [4]. This change can have consequences on a proper result interpretation. Biological material is sent for analysis for diverse purpose. During regular control check of physiological state of organism, disease diagnosis and prognosis in clinical settings and basic research purpose of investigating genetic, biochemical and molecular interactions of key molecules in health and disease [5, 6]. The most commonly used biological material for analysis is Blood by phlebotomy [7]. Blood contains blood cells (Erythrocyte_ER, Leukocyte_LE and Platelet_Pt) and blood fluid, Plasma (PL) [7]. Blood plasma is gained by adding anticoagulants [8].

Cholinesterase (ChE) enzymes are promising blood biomarkers for disease diagnosis, prognosis and treatment [9]. Based on substrate and inhibitor specificity cholinesterases are classified as Acetylcholinesterase (AChE) and Butyrylcholinesterase (BChE) [10]. AChE, true ChE, is found in red blood cells, while BChE, pseudocholinesterase, is found in plasma or serum [11]. Natural and synthetically produced inhibitors of enzyme AChE activity are used for disease treatment [12]. Storage time is one of the factors influencing biomarker activity, among which is enzyme AChE. Various articles have been published regarding this topic. However, the most common research was done on diverse animal models [13-15]. Few studies have been conducted on human samples with maximum 1 month of storage on limited types of blood constituents [16-19]

Moreover, water intake as an environmental factor has impact on blood biomarker range. Water intake causes change in active site of enzyme AChE [20]. There is no information regarding the influence of different water intake amount and storage time on protein concentration, enzyme AChE activity change and inhibitor efficacy of inhibitor BW284c51, GUK-987 and GDK-510 in human blood from the literature.

Therefore, current study done as a part of PhD thesis is an attempt to contribute in gaining new knowledge regarding the influence of water intake before sampling and 92 days of storage at −20°C in the freezer on enzyme AChE activity from Plasma and Erythrocyte samples. This study connects the previous paper from PhD student Jovicic S, form faculty of Biology, University of Belgrade, Serbia regarding the influence of water intake on enzyme AChE activity. Samples conducted from this study have been frozen. Research task is defined by the fact that enzyme AChE activity is subjected to change due to storage time. Since enzyme AChE is a key biomarker for human disorders, tested hypothesis postulates that water intake before sampling and storage time influence Erythrocyte and Plasma level of protein concentration, enzyme AChE activity and inhibitor efficacy in healthy subjects.

In order to test the working hypothesis, the objective of the paper was to examine effect of total **92 days** of blood storage on in vitro **protein concentration, basal enzyme AChE activity and inhibition efficacy** of inhibitors **BW284c51**, 0.01mM and **GUK-987**, 0.1mM. This paper investigate how water intake level and storage time affect protein concentration, enzyme AChE activation/inhibition in healthy subjects from ER (0 day vs 90^th^, 91^st^ day) and PL samples (0 day vs 92^nd^ day). Moreover, the objective of the paper is expanded to investigate the most efficient concentration of inhibition for inhibitor GUK-987 and GDK-510 for further clinical studies.

## 2. Material and methods

### 2.1 Material

#### 2.1.1 Human biological material

Experiments were performed according to **ethical standards** and with written consent of the blood donors. The study was given ethical permission from the National Medical Ethics Committee (number 82/07/14). Experimental part of the PhD thesis was done at the Biotechnical Faculty, University of Ljubljana, Slovenia, within the project **J5-7098** and **CEEPUS free mover PhD student program mobility** with the University of Belgrade, Serbia.

Total number of samples consisted of **11 healthy participants, (T=6, C=5)**. Storage samples differ based on blood sample type (PL and ER), amount of water intake (Test_T and Control_C group) and inhibitor (BW284c51/GUK-987). Study evaluated the influence of storage time (0, 90^th^, 91^st^ and 92^nd^ day) at −20°C in the freezer on protein concentration, enzyme AChE activity and inhibitor efficacy from samples of healthy male and female individuals.

### 2.2 Methods

#### 2.2.1 Sampling of biological material

Whole blood (WB) samples were collected from healthy individuals by venepuncture using needles of 21 diameter (70mm length, 0.4 mm inner diameter, Microlance, Becton Dicinson, Franclin Lakes, NJ, USA) and stored in four 2.7mL vacutainer containing 270 µL trisodium citrate (0.109 mol / L) as anticoagulant.)

Blood elements, plasma and erythrocyte membrane, were isolated from the subject biological material. Protein concentration was assessed on microtiter plate. The measurement of cholinesterase activity in plasma and erythrocyte samples was accomplished using the Elman et all method (1961).

#### 2.2.2 Fractioning of biological material

##### a) Plasma isolation

100µL of whole blood was sampled into the ependorf tube to measure cholinesterase activity. In order to separate cells from plasma, blood was centrifuged at 1550 g, 37 ° C, for 20 minutes in a centric 200/R centrifuge (Domel doo, Železniki, Slovenia). After a first centrifugation, the upper plasma fraction was separated in a centrifuge tube (1.5 mL). Plasma was placed in Ependorf tubes. Leukocyte sheath was removed from red blood cells before and after centrifugation.

The red blood cells were washed twice in phosphate-citrate buffer solution (PBS citrate buffer; 137mM NaCL, 2.7 mM KCL, 7.8 mM Na2HPO4 * 2H2O, 1.5 mM KH2PO4, containing 10.9 mM trisodium citrate; pH 7.4) for 5 min. at 1550 g, 37°C; Centric 200/R centrifuges (Domel doo, Železniki, Slovenia).

Note1: all plasma or part of the buffer (supernatant) was removed and replaced with 1 mL of fresh PBS citrate buffer; after the final second centrifugation, the supernatant was removed and the red blood cells were stored in the same vial.

Note2: As a preliminary test, the leukocyte sheath was stored in a 2.7mL vial without the presence of anticoagulants and analyzed without washing or further processing.

For the next experiment: The leukocyte sheath was pipette using 1mL wider extensions, while the polysaccharide gradient was used to isolate the monocytes. The procedure used did not provide a pure white blood cell fraction as the plasma and red blood cells remained in the fraction. The results will not be considered in the analysis.

##### b) Erythrocyte membrane isolation

During erythrocyte membrane fractionation, 500 µL of whole blood was collected in 1.5mL ependorf tube using 100 µL extensions (note: it is best to use a larger extension than 1mL during pipetting, in order to avoid the destruction of red blood cells). Centrifuge at 200 g (= 1440 rpm, Železniki Technique, 322A) for 5 min, at 4°C to remove plasma. About 200 µL of blood plasma was removed and the remaining suspension was washed 3 times with cold 1X PBS citrate buffer (10mM PBS citrate buffer; 137 mM NaCL, 2.7 mM KCl, 7.8 mM Na2HPO4 • 2H2O, 1.5 mM KH2PO4 containing 10.9mM trisodium citrate; pH 7.4). The erythrocytes were resuspended in hypotonic PBS citrate buffer with 5mM Tris HCL containing 1mM EDTA pH 7.4 (about 1000 µL fresh buffer added after washing) and left at 10°C overnight. The erythrocytes were centrifuged 7-8 times in hypotonic PBS citrate buffer at 9000 rpm for 10 min at 4°C to obtain erythrocyte membranes (ER spirit) at the base of the ependorf. The erythrocyte membranes thus prepared (after loss of hemoglobin) were treated 3X for 30 sec with Triton X-100 detergent (0.1%, prepared in 25mM phosphate buffer, pH 7.4) to dissolve the membrane-bound enzyme, acetylcholine esterase (AcetylCholin esterase-AChE). The homogenate was centrifuged at 10,000 g for 10 min at 4-6 °C and the pure supernatant was used for AChE analysis.

#### 2.2.3 Biochemical methodology

##### a) Measurement of protein concentration

To measure protein absorption on a microtiter plate, we have 3 types of reaction: sample reaction, negative control reaction, and standard curve (bovine serum albumin-BSA) dilution of 2mg / ml, 1mg / ml, 0.5mg / ml, 0.25mg / ml, 0.125mg / ml, 0.0625 mg / ml). We did triplicates for each reaction. Depending on the type of reaction in each well, we had: 20 µL of sample / buffer / BSA dilution and 200 µL of a mixture of reagent A and reagent B in a ratio of 1:50). The absorbance was measured at **550nm** using a spectrophotometer (BioTek, Cytation 3, Bad Friedrichshall, Germany) for one cycle (30 sec). Based on the absorbance values obtained, the protein concentration of each sample was calculated using a Microsoft excel 2007 program.

Technical view1: To prepare a dilution solution, equation C1=(C2xV2)/V1 is used to calculate stock solution. Upon gaining absorbance values of triplicate, mean value was calculated in Microsoft Excel 2007. Blank value is subtracted from the average. Standard curve of linear regression with equation is assessed in the form y=aX + b and R2 value, where a is line slope; b is intercept, and R is correlation coefficient. R2 value closer to 1 indicated a good fit of the standard curve and linear regression. Protein concentration [mg/ml] is calculated, taking into the account dilutions for each sample.

##### b) Measuring cholinesterase activity

To measure cholinesterase activity on a microtiter plate, we made 3 types of reaction, induced reaction, inhibited reaction, and negative control reaction. We did triplicates for each reaction. Depending on the type of reaction in each well for the endogenous/induced/inhibited reaction, we have 100 µL of diluted sample, 100 µL of Elman reagent (DTNB in 250mM KP buffer, pH 7.4) without/with acetylthiocholine chloride substrate (final concentration of 1mM) and 20 µL of KP buffer/inhibitor (KP buffer/BW284c51; GUK-987/GDK-510). In the negative control reaction, we have 120 µL K-P buffer and 100 µL Elman’s substrate reagents. Absorbance was measured at **420nm** using a spectrophotometer (BioTek, Cytation 3, Bad Friedrichshall, Germany) for 40 cycles (30 sec interval per cycle, 20min). Using the protein concentration and mass of each sample as well as the difference in absorbance and reaction rate over the selected time frame (linear absorbance selection/time cycle), the enzymatic activity of each sample is calculated using the Microsoft Excel 2007 program.

Technical view 2: To calculate enzyme activity, equation c=n/V is used, where C is concentration of the solution (mol/L), n is moles of substance being dissolved (moles of solute) and V is volume of the solution in liters (L). Mass of protein is expressed in **mg/ml unit**. Change of absorbance in min is calculated and concentration in minute is assessed (mM) utilizing Lamber Beer Law. Spead of reaction is calculated as change of concentration in minute multiplied by Total volume, and is expressed in **nmol/min**. Final cholinesterase activity is calculated dividng reaction speed and protein mass, and is expressed in **nmol/min/mg** protein.

## 3. Statistical analysis

Data analysis was performed by **IBM SPSS software** version 23.0 for statistical analysis. **Descriptive statistics, Pearson correlation, Linear Stepwise regression and Paired T test, One way ANOVA** was assessed. Results are expressed as **mean +-SD** and **p value**. Statistical significant correlation was assumed when **p<0.05**.

## 4. Results

### 4.1 Participants information

Difference among participants is based on amount of drinking water, gender, protein concentration, AChE activity and inhibitor efficacy in Erythrocyte and Plasma samples of healthy individuals. Effect of storage time at −20°C in freezer is measured at 0, 90^th^, 91^st^ and 92^nd^ day.

**Descriptive statistics** of the participants include mean and standard deviation value for followed characteristics of **protein concentration, basal enzyme AChE activity and inhibitor efficacy** between Gender (Male and Female) and Sample group (Test and Control).Protein concentration is expressed in **mg/ml**, while enzyme activity is expressed as **nmol/min/mg**. All data are parametric, with normal, Gaussian curve distribution. Further statistical analysis employed **parametric** statistical test. Participant characteristics are presented in **Table 1. Table 1** show characteristics of the frozen samples, included in the study based on gender and participants group in PL and ER blood samples.

**Table 1.** Participant’s characteristics of protein concentration, basal AChE activity, inhibitor efficacy based on gender and sample group

| Table 1. Frozen samples characteristics, 0, 90<sup>th</sup>, 91<sup>th</sup>, 92<sup>nd</sup> and 93<sup>rd</sup> day |  |  | Group Sample |  |  |  |
| --- | --- | --- | --- | --- | --- | --- |
|  |  |  | C |  | T |  |
|  |  |  | Mean | Standard Deviation | Mean | Standard Deviation |
| PL_7.6_(0 day)_Protein concentration | Gender | F | 130.73 | 65.51 | 112.92 | 35.92 |
|  |  | M | 97.81 | 35 | 111.30 | 35 |
| PL_7.6_(92 day)_Protein concentration | Gender | F | 43.84 | 11.26 | 55.06 | 9.36 |
|  |  | M | 39.96 | 9 | 43.31 | 9 |
| PL_7.6_(93 day)_Protein concentration | Gender | F | 41.25 | 11.44 | 52.40 | 8.20 |
|  |  | M | 37.00 | 9 | 41.00 | 7 |
| ER_9.6_(0 day)_protein concnetration | Gender | F | 34.66 | 6.95 | 33.21 | 6.87 |
|  |  | M | 28.87 | 7 | 29.36 | 6 |
| ER_9.6_(90 day)_Protein cocncentration | Gender | F | 24.93 | 0.71 | 26.93 | 4.72 |
|  |  | M | 23.34 | 0.6 | 25.13 | 4.1 |
| ER_9.6_(91 day)_Protein cocncentration | Gender | F | 25.50 | 4.36 | 24.20 | 5.07 |
|  |  | M | 26.00 | 4.5 | 27.00 | 5.5 |
| PL_hi_I_Basal_AChE_(7.6)_0day | Gender | F | 0.07 | 0.01 | 0.05 | 0.02 |
|  |  | M | 0.08 | 0.01 | 0.03 | 0.01 |
| PL_hi_I_Basal_AChE(7.6vs8.9)_92day | Gender | F | 0.02 | 0.00 | 0.01 | 0.00 |
|  |  | M | 0.02 | 0.00 | 0.01 | 0.00 |
| ER_hi_I_Basal_AChE(9.6)_0 day | Gender | F | 0.63 | 0.09 | 0.49 | 0.10 |
|  |  | M | 0.74 | 0.07 | 0.54 | 0.2 |
| ER_hi_I_Basal_AChE_(8.9)_90day | Gender | F | 0.24 | 0.03 | 0.20 | 0.05 |
|  |  | M | 0.19 | 0.01 | 0.18 | 0.04 |
| ER_hi_I_Basal_AChE_(9.9)_91day | Gender | F | 0.20 | 0.02 | 0.17 | 0.04 |
|  |  | M | 0.18 | 0.01 | 0.14 | 0.03 |
| PL_hi_I_inh.BW285c51_0.01mM_AChE_(7.6)_0 day | Gender | F | 0.05 | 0.01 | 0.03 | 0.01 |
|  |  | M | 0.06 | 0.01 | 0.02 | 0.01 |
| PL_hi_I_inh.BW285c51_0.01mM_(7.6 vs 8.9)_92day | Gender | F | 0.05 | 0.01 | 0.03 | 0.01 |
|  |  | M | 0.06 | 0.01 | 0.02 | 0.01 |
| ER_hi_I_inh.BW285c51_0.01mM_(9.6)_0day | Gender | F | 0.00 | 0.00 | 0.00 | 0.00 |
|  |  | M | 0.00 | 0.00 | 0.00 | 0.00 |
| ER_hi_I_inh.BW285c51_0.01mM_(8.9)_90day | Gender | F | 0.00 | 0.00 | 0.00 | 0.00 |
|  |  | M | 0.00 | 0.00 | 0.00 | 0.00 |
| PL_hi_I_inh.GUK 987_0.1mM_(7.6vs9.9)_93 day | Gender | F | 0.00 | 0.00 | 0.00 | 0.00 |
|  |  | M | 0.00 | 0.00 | 0.00 | 0.00 |
| ER_hi_I_inh.GUK-987_0.1mM_(9.6)_0 day | Gender | F | 0.30 | 0.05 | 0.25 | 0.05 |
|  |  | M | 0.34 | 0.06 | 0.27 | 0.06 |
| ER_hi_I_inh.GUK_0.1mM_(9.9)_91day | Gender | F | 0.01 | 0.00 | 0.01 | 0.00 |
|  |  | M | 0.00 | 0.00 | 0.01 | 0.00 |

**Gender and sample group** differs based on amount of drinking water. On average, control group drank 2141.67 ml while Test group drank 1656.25 ml. Moreover, Female drank 230 ml more in comparing to males who drank in total 1950 ml of water. **Table 1**. illustrates the mean value and standard deviation of protein concentration, basal AChE activity and inhibitor efficacy of inhibitors GUK-987 and BW284c51 in Plasma and Erythrocyte samples. Descriptive statistic showed that **protein concentration** and **basal AChE activity** varies depending on storage time, sample type (PL/ER) and group (T and C), while **inhibitor efficacy** in the same group remains the same. Inhibitor 100% efficacy is presented in PL samples with inhibitor GUK-987 and in ER samples for inhibitor BW284c51. Results indicate that storage time influence change in concentration of protein, basal AChE activity and inhibitor efficacy.

**Protein concentration is** clearly the highest in Female Control group of Plasma samples and the lowest in ER samples. Graphical representation of protein concentration, enzyme activity and inhibition efficacy is presented in the next **graphs** (1-4). **Test group** is presented in participant samples number 1, 2, 5, 6, 7, 9, while **Control group** is presented in participant samples number 3, 4, 8, 10, 11.

**Graph1.**
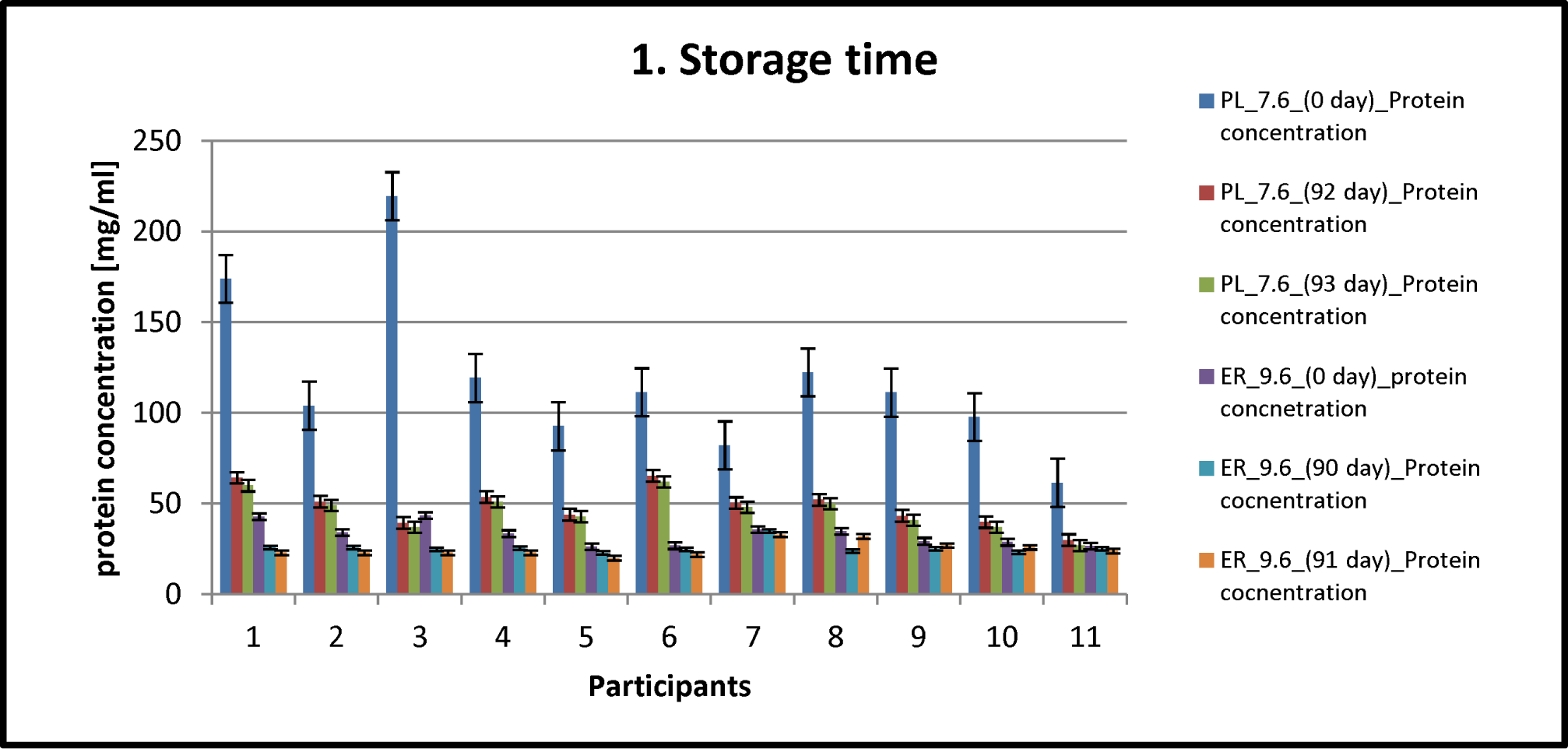
Protein concentration with error bars with standard errors in Plasma and Whole blood samples after storage of 0, 90^th^, 92^nd^ and 93^rd^ days on −20°C degrees in the refrigerator.

**Graph2.**
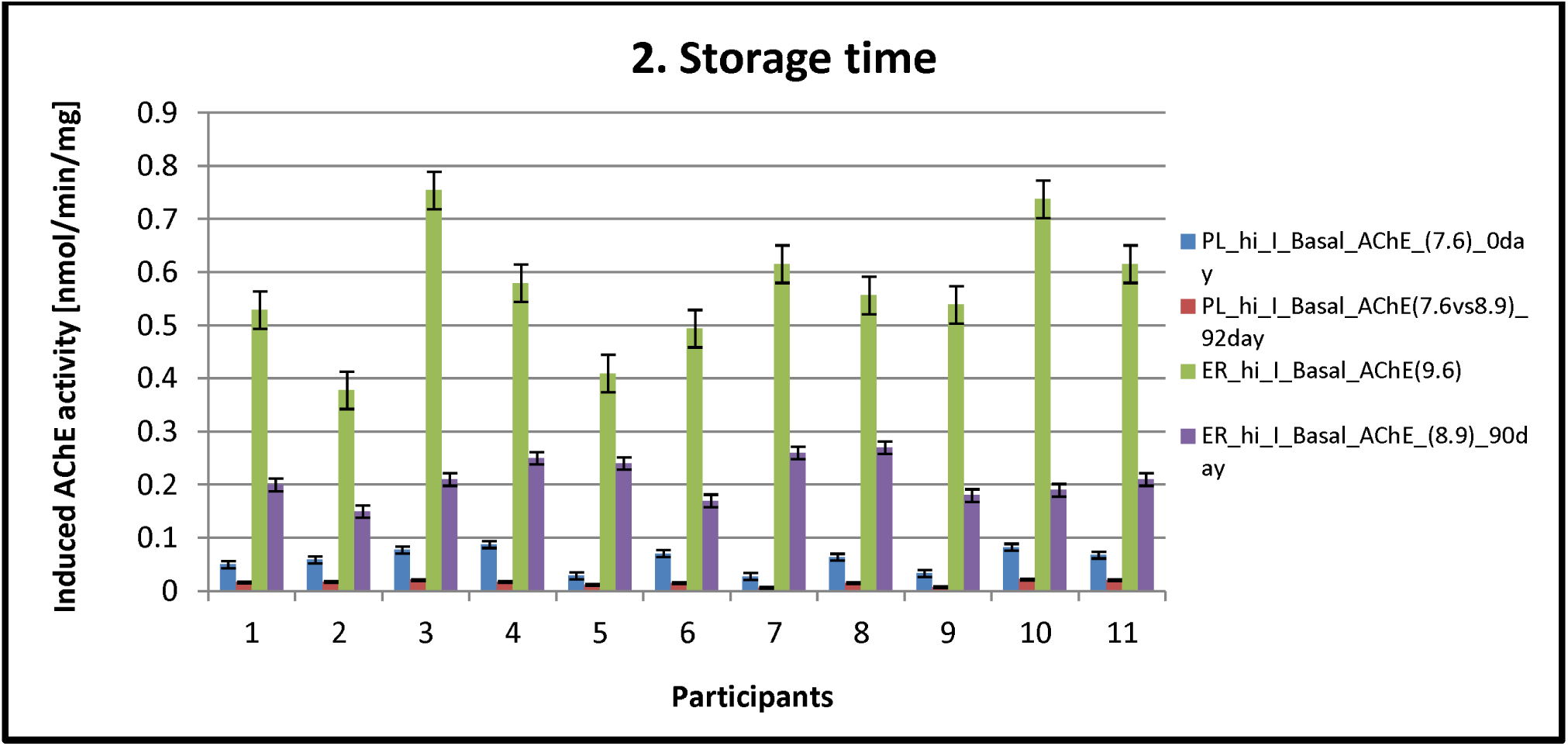
Induced enzyme AChE activity with error bars with standard errors in Plasma and Whole blood samples after storage of 0, 90^th^, 92^nd^ and 93^rd^ days on −20°C degrees in the refrigerator.

**Graph3.1.**
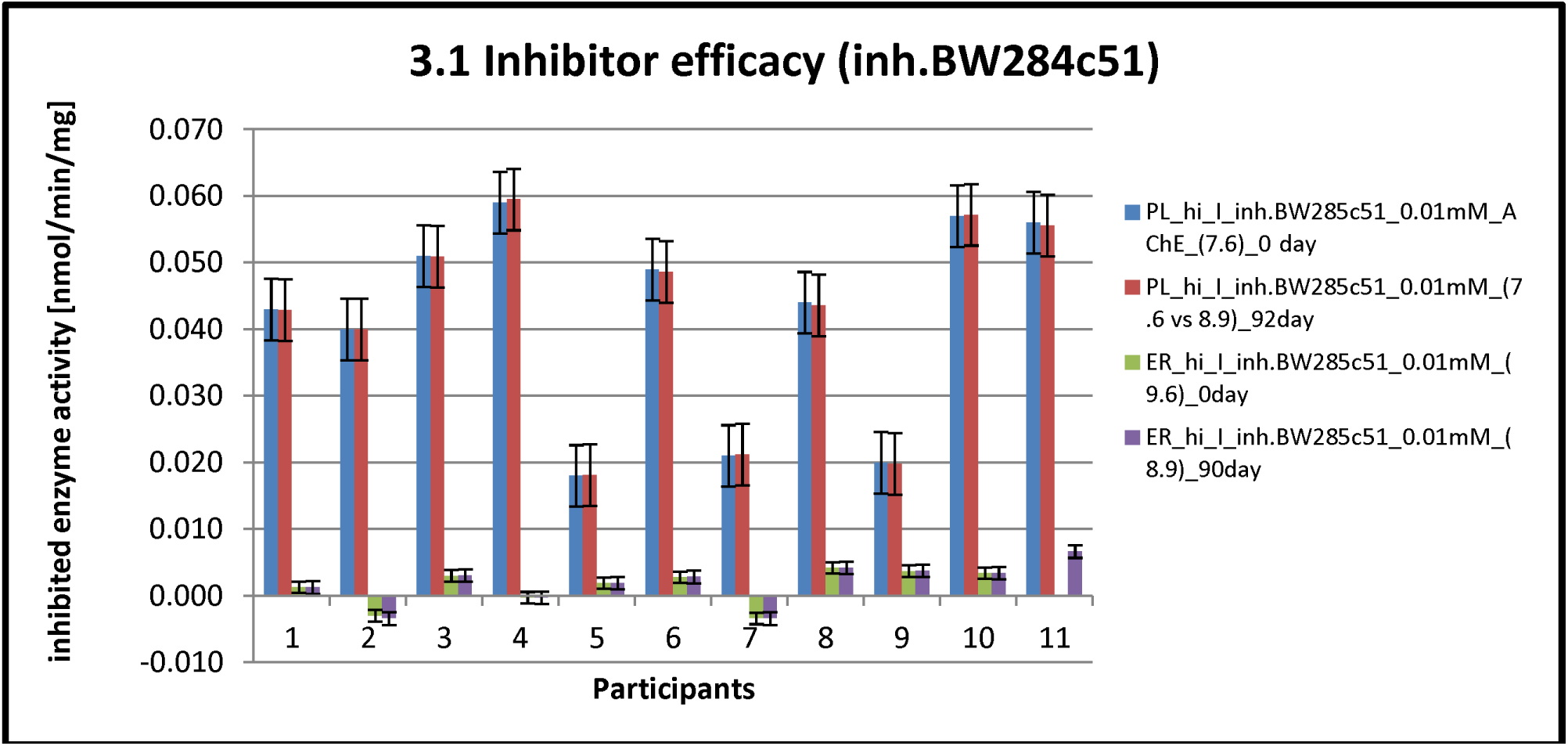
Inhibited enzyme AChE activity (inhibitor BW284c51, 0.01mM) with error bars with standard errors in Plasma and Whole blood samples after storage of 0, 90^th^ and 92^nd^ days on −20°C degrees in the refrigerator.

**Graph3.2.**
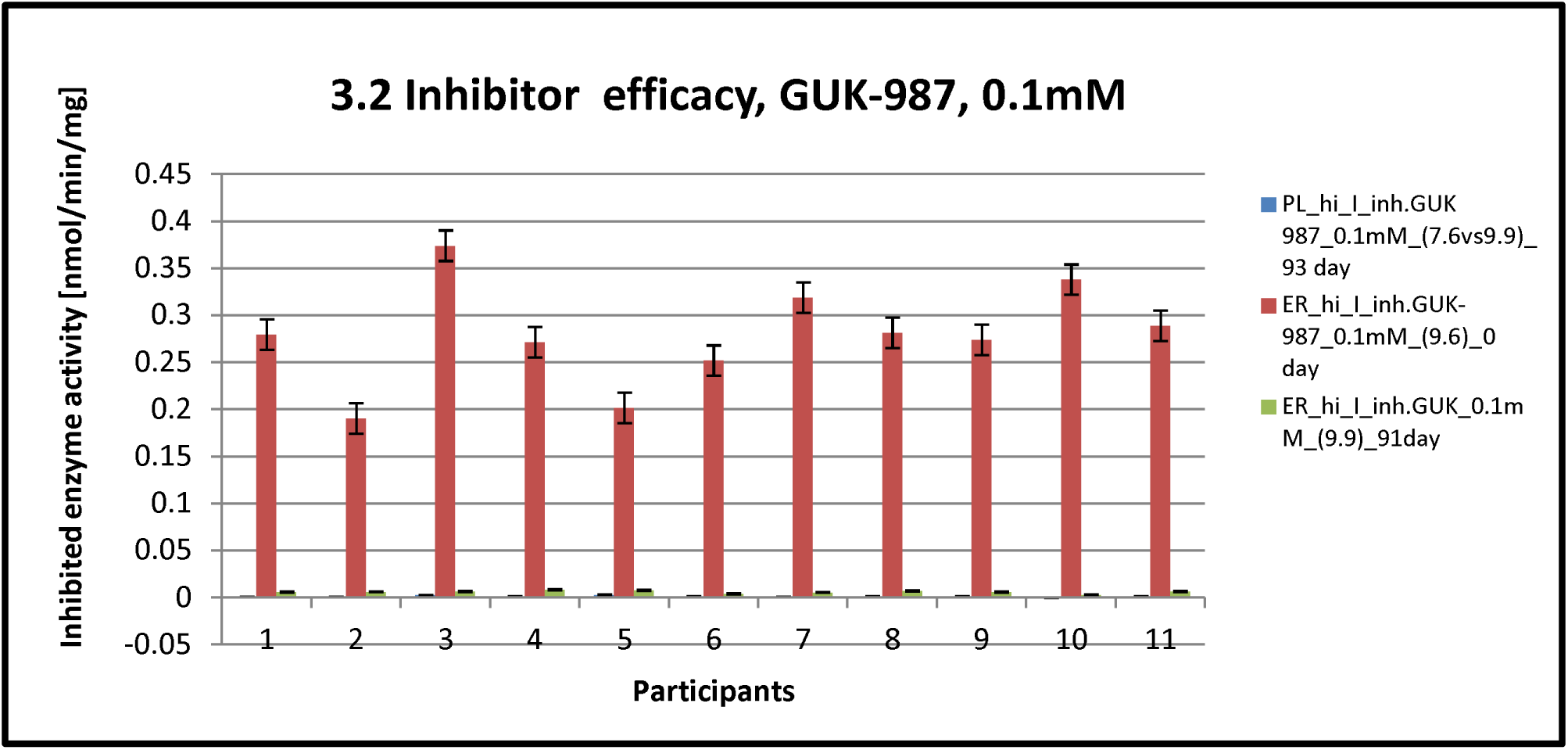
Inhibited enzyme AChE activity (inhibitor GUK-987, 0.1mM) with error bars with standard errors in Plasma and Whole blood samples after storage of 0, 91^th^ and 93^rd^ days on −20°C degrees in the refrigerator.

### 4.2 Effect of **water intake** and **storage time** on protein concentration, enzyme AChE activity and inhibitor efficacy

In order to investigate the effect of storage time on protein concentration, basal AChE activity and inhibitor efficacy, this research study employs usage of Descriptive statistics, Pearson correlation, Regression analysis (linear, enter, logistic), Paired T test and One way ANOVA).

#### 4.2.1 Descriptive statistics

**Protein concentration is** clearly the highest in Female Control group of Plasma samples and the lowest in ER samples. <u>Plasma samples</u> protein concentration in **control groups** for Females/Males decreased dramatically 2.98/2.44 times for 92 days of storage to 3.16/2.64 times for 93 days of storage. Difference between 92^nd^ and 93^rd^ day is minor, 1.06/1.08 times for male and female. **Test group** shows concomitant decrease for Females/Males 2.05/2.57 times for 92 days of storage to 2.15/2.71 times for 93 days. The same decrease for Female/Male, 1.05, is seen between 92^nd^ and 93^rd^ day. Similar fall in protein concentration is noticeable in Erythrocyte samples. Decrease in **Control group** is seen at a slower pace in Males/Females from about 1.34/1.23 for 92 days of storage and 1.35/1.11 for 93 days of storage. Protein concentration difference fall in Female/Male between 92^nd^ and 93^rd^ day is 0.97/0.89 times. In contrast, **Test group**, indicated decrease of protein concentration after 92/93 days of storage for 1.23/1.37 times in Females and 1.16/1.09 time for Males. Difference between 92^nd^ and 93^rd^ day is in Female/Male dropped for 1.11/0.93 times, from approximately the same level as in Plasma samples.

Comparing protein concentration in Plasma and Erythrocytes samples, results illustrates significantly higher decrease in plasma samples.

**Basal AChE activity** shows changes in control and test group. Erythrocyte samples demonstrated the highest basal AChE activity. The most striking feature of the Table 1 is the drop of basal AChE activity after 91 days of storage. In **control group** it is shown a dramatic decrease of activity in **female** participants from 63% at 0 day to 24% and 20% for 90^th^ and 91^st^ day and **male participants** from 74% at day 0 to 19% and 18% for 90^th^ and 91^st^ day of storage at −20°C. By contract, based on the above information, it is predicted that difference between 90^th^ and 91^st^ day of storage be lover 4% and 1% in female and male participants. **Test group** show the most significant fall in basal AChE activity after 91 days of storage. Total decrease of Basal AChE activity for 91 days of storage is 17% and 14% in Female/Male participants. Starting activity from day 0 was 49% and 54% retrospectively for Female/Male participants. This activity drops in female/male participants after 90 days of storage to 20% and 18%. Plasma samples showed decrease of basal AChE activity of **Control group** from 7% to 2% in female participants and 8% to 2% in male participants at 0 and 92^nd^ day of storage at −20°C. Moreover, **Test group** had similar fall, from 5% to 1% in Females and 3% to 1% in Males at 0 and 92^nd^ day of storage.

It is clear that the **Male participants** have the highest basal AChE activity in **ER samples** at day 0, and decrease of activity after 91 days. **Plasma samples** showed the same conclusion, with different starting and ending enzyme AChE activity. However, ER samples have the highest starting basal AChE in comparing to PL samples, 74% in ER and 7% in PL samples. There is 10 times higher basal AChE activity in ER samples of males participants

**Inhibitor efficacy** remained the same for inhibitor BW284c51 0.01mM for **PL and ER samples** during 0 and 90 days of storage at −20°C. The most efficient inhibition of enzyme AChE with inhibitor BW284c51 0.01mM is in ER samples (100% of inhibited AChE activity) in both control and test group. In Plasma samples, test group showed the highest efficacy in comparing to control group. Females and males from **control group** showed decrease of enzyme AChE activity to 5%/6% at 0 and 92^nd^ day of storage. Test group decreased enzyme AChE activity to 3% and 2% in female and males for 0 and 92^nd^ day of storage.

The most striking efficacy inhibition of enzyme AChE effect is seen with inhibitor GUK-987, 0.1mM in PL samples at 0 and 93^rd^ day **in control and test group**. In ER samples, inhibitor GUK 987 efficacy increased after 91 day of storage, for 1% and 0% in Females/Males in comparing to the starting efficacy of 30% and 34% in female/males before storage in control group. Test group showed inhibition from 25%/27% to 1% at 0 and 91^st^ day of storage in female and male participants.

Results from descriptive statistics indicated mean difference between investigated variables and data normality. In order to see if there is statistical significant difference between them, **parametric test** were used.

#### 4.2.2 Pearson correlation

Pearson correlation showed statistical significant linear correlation (p<0.05*, p<0.01**) for the investigated variables.

**Protein concentration** from PL samples showed correlation in **Test group** at *day 0* with control group, within Test group at 92^nd^ and 93^rd^ *day*, and with inhibitor BW284c51 at 0/92 day. Test group at 93^rd^ day is correlated with protein concentration at 92^nd^ day, inhibitor BW284c51 at 0/92 day. **Protein concentration** from PL samples showed correlation in **Control group** at 0 *day* with PL test group and ER samples control group at 0 day. Control group at 92^nd^ *day* of storage showed correlation to 93^rd^ day.

**Basal AChE enzyme activity** from PL samples showed correlation in **Test group** at 0 *day* with inhib.BW284c51 at 0/92 day, control group for basal AChE activity at 0 day. Test group at 92^nd^ day with inhibitor BW284c51 at 0/92 day. **Basal AChE enzyme activity** from PL samples showed correlation in **Control group** at *day 0* with 90^th^ day, ER basal AChE activity at 90^th^/91^st^ day. Control group at 92^nd^ *day* of storage showed correlation to basal AchE activity in ER samples at 90nd day.

**Inhibitor BW284c51 efficacy** from PL samples showed correlation in **Test group** at 0 day with protein concentration at 92^nd^/93^rd^ day, Basal AChE activity at 0/92^nd^ day, inhibitor BW284c51 at

92^nd^ day. Test group at 92^nd^ day showed correlation with protein concentration at 92^nd^/93^rd^ day, basal AChE activity at 0/92^nd^ day, inhibitor BW284c51 at 0 day. **Inhibitor BW284c51 efficacy** from PL samples showed correlation in **Control group** with 0/92^nd^ day.

**Protein concentration** from ER samples showed correlation in **Test group** at *90*^*th*^ *and 91*^*st*^ *day*.

**Basal AChE enzyme activity** from ER samples showed correlation in **Test group** at 0 day with inhibitor.GUK 987. Test group at 90 day showed correlation with basal AChE activity at 91^st^ day while test group at 91^st^ day showed correlation with basal AChE activity at 0 day. **ER sample Basal AChE enzyme activity** from ER samples showed correlation in **Control group** at 0 day with ER_samples inhibitor GUK at 0 day. **Control group** at 90 *day* showed correlation with 92^nd^ day.

**Inhibitor BW284c51 efficacy** from ER samples showed correlation in Test group at day 0 with 90^th^ day. **Inhibitor BW284c51 efficacy** from ER samples showed correlation in Control group at 0 and 90^th^ day.

#### 4.2.3 Regression

##### 4.2.3.1 Linear regression

**Linear stepwise regression** showed linear relationship between variables (protein concentration, basal AChE activity and inhibitor efficacy) in test and control groups. Protein concentration variable in **Test group** showed correlation of PL_samples during 92^nd^ and 93^rd^ day, ER_samples during 90^th^ and 91^st^ day. **Control group** showed correlation of PL samples during 92^nd^ and 93^rd^ day, PL and ER samples at day 0. **Between Test and Control group** correlation is shown of PL samples for day 0. Basal AChE enzyme activity **variable in Test group** showed correlation of ER_samples during 90^th^ and 91^st^ day, ER_samples at 0 day between basal AChE and inh.GUK. **Control group** showed correlation of PL_samples during 90^th^ and 92^nd^, ER_samples during 90 and 92^nd^ day, ER_samples at 0 day between basal AChE and inh.GUK. Inhibitor efficacy of inhibitor BW284c51 variables in **Test group** showed correlation of PL_samples during 0 and 92^nd^ day, ER_samples during 0 and 90^th^ day. **Control group** showed correlation of PL_samples during 0 and 92^nd^ day, ER_samples during 0 and 90^th^ day. Association is confirmed with cross validation. Age is not statistical significant variable in our case, so the graph is made without it.

###### Graphical representation of linear regression model for Control and test group is shown in Graphs 4.1-4.12

**Graph 4.1.**
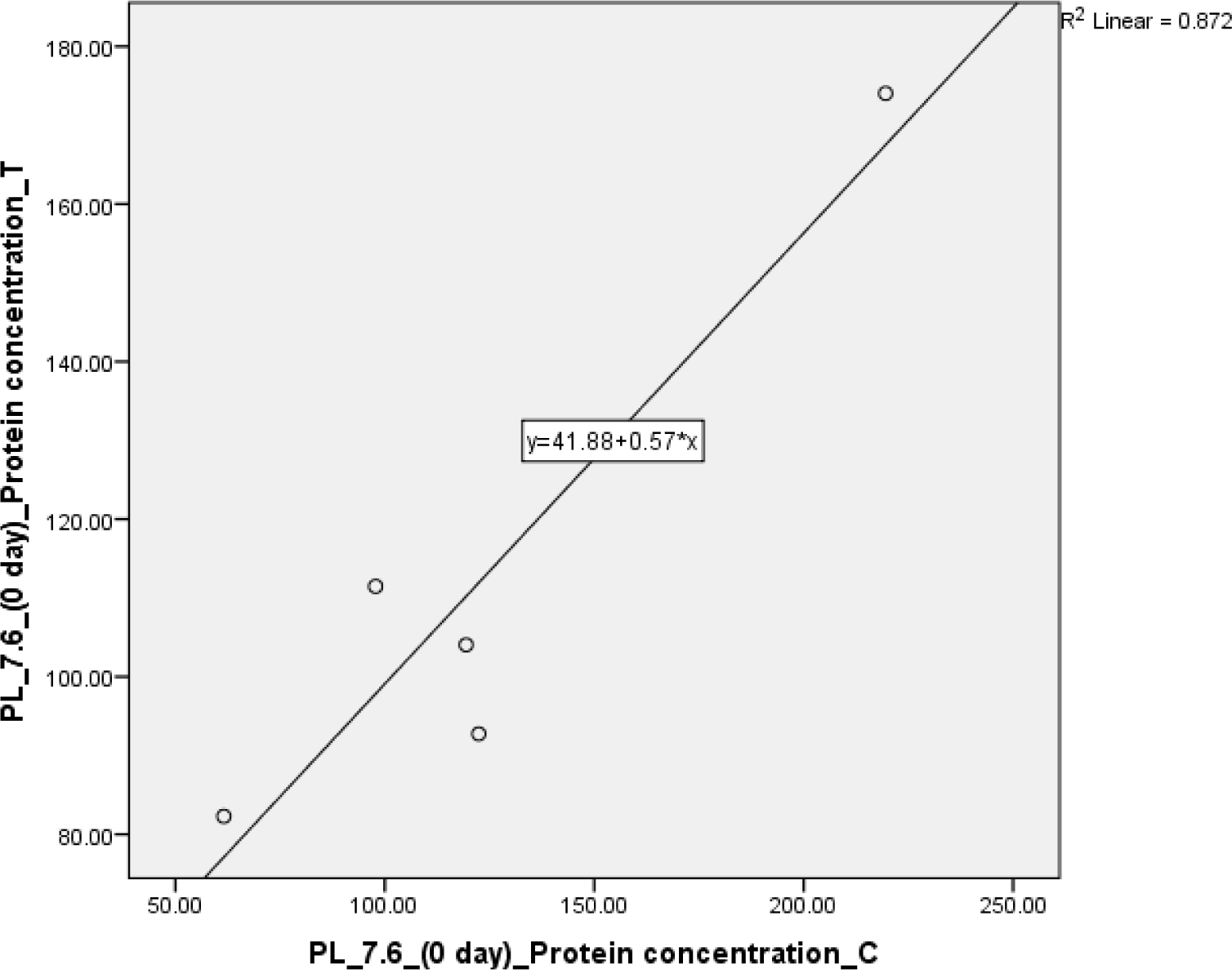
Linear stepwise regression correlation for Protein concentration before freezing (0 day) in PL samples between Test and Control group (R2=0.872), which is confirmed with cross validation methodology for 80%samples (p<0.05, R=0.934*)

**Graph 4.2.**
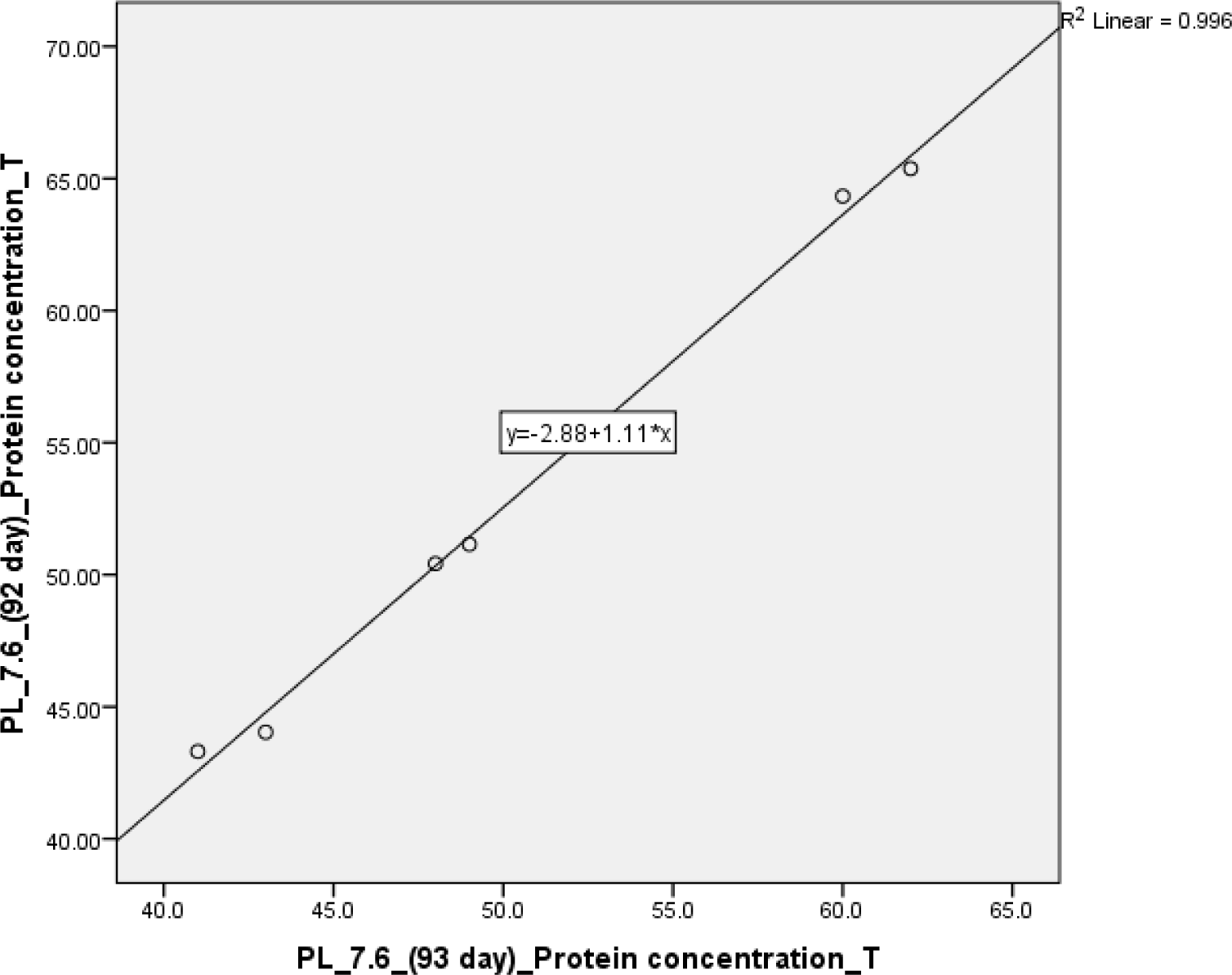
Linear stepwise regression correlation for Protein concentration at 92^nd^ and 93^rd^ day of freezing, in PL samples of Test group (R2=0.996), which is confirmed with cross validation methodology for 80%samples (p<0.01, R=0.998**)

**Graph 4.3.**
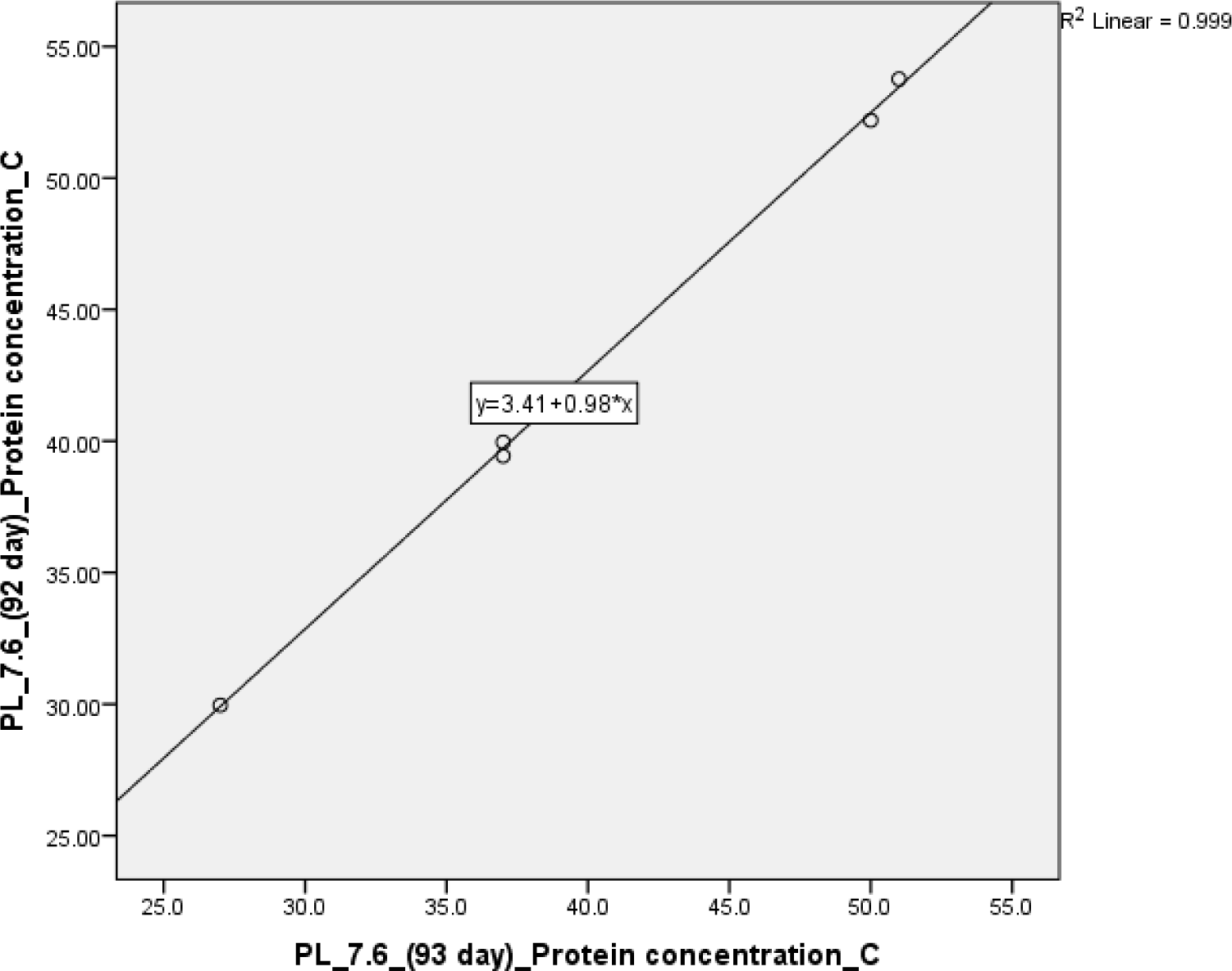
Linear stepwise regression correlation for Protein concentration at 92^nd^ and 93^rd^ day of freezing, in PL samples of Test group (R2=0.999), which is confirmed with cross validation methodology for 80%samples (p<0.01, R=1.000**)

**Graph 4.4.**
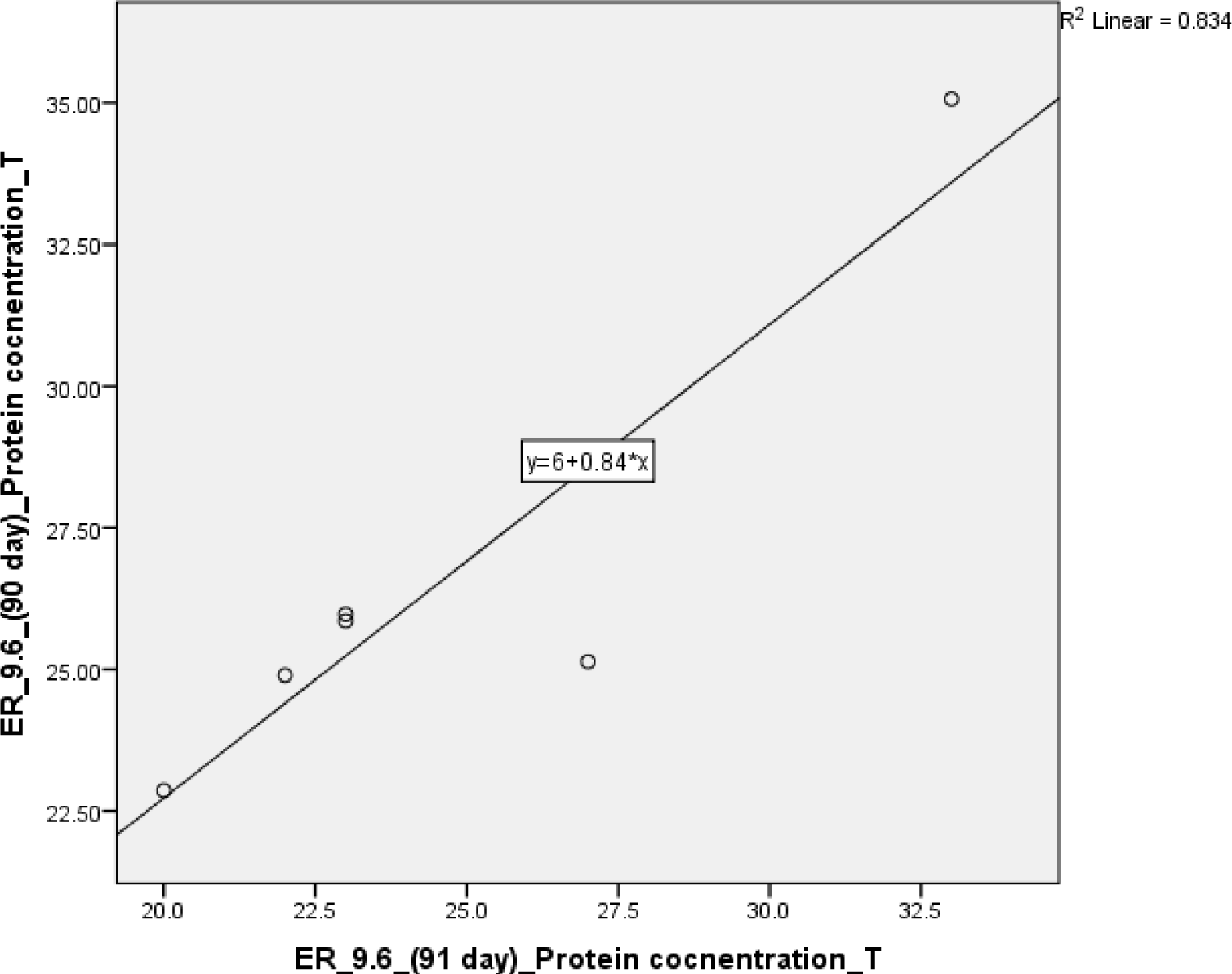
Linear stepwise regression correlation for Protein concentration at 90^th^ and 91^st^ day of freezing, in ER samples of Test group (R2=0.834) confirmed with cross validation methodology for 80%samples (p<0.01, R=0.913*)

**Graph 4.5.**
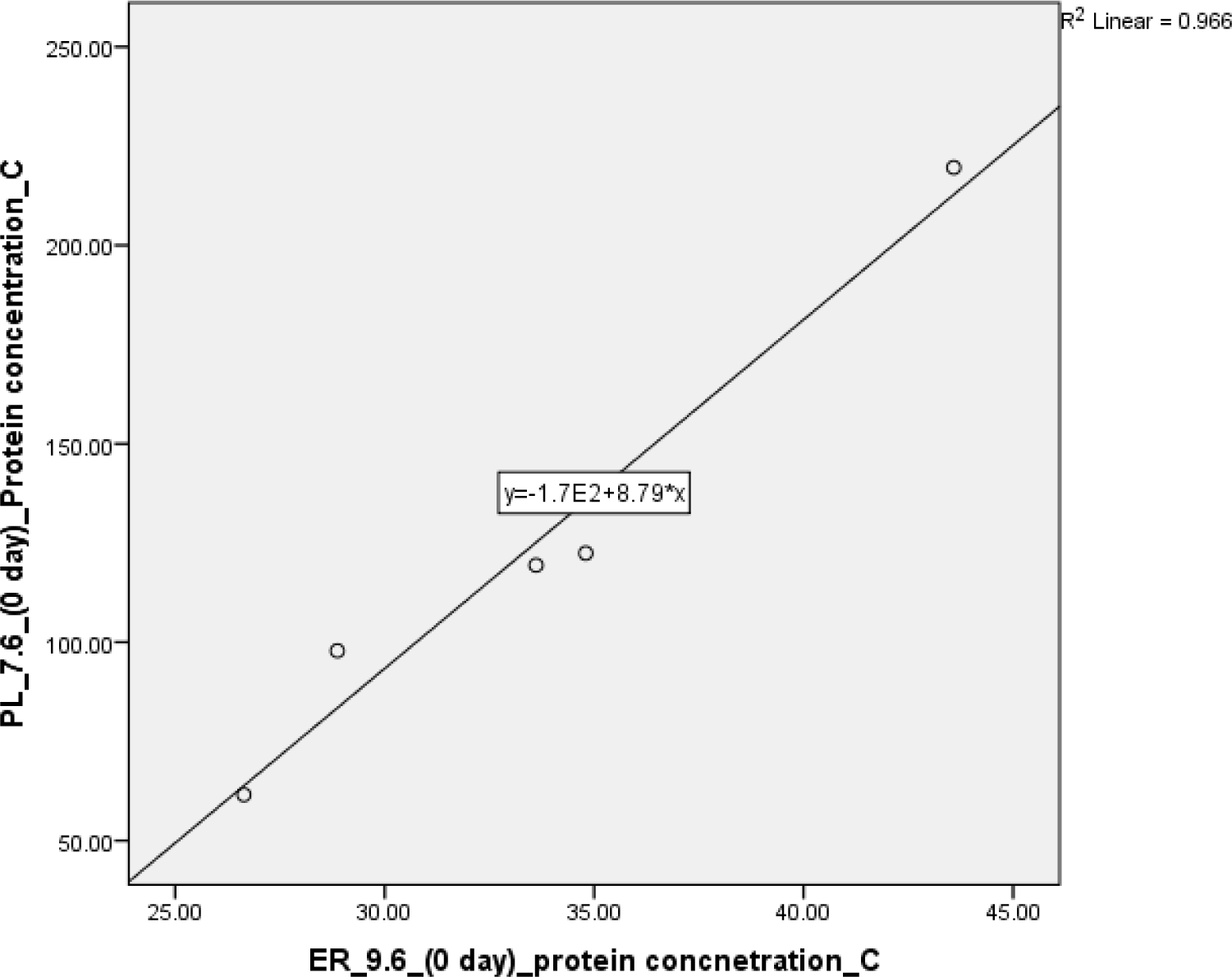
Linear stepwise regression correlation for Protein concentration at 0 day of freezing, in PL and ER samples of Control group (R2=0.966) confirmed with cross validation methodology for 80%samples (p<0.01, R=0.983**)

**Graph 4.6.**
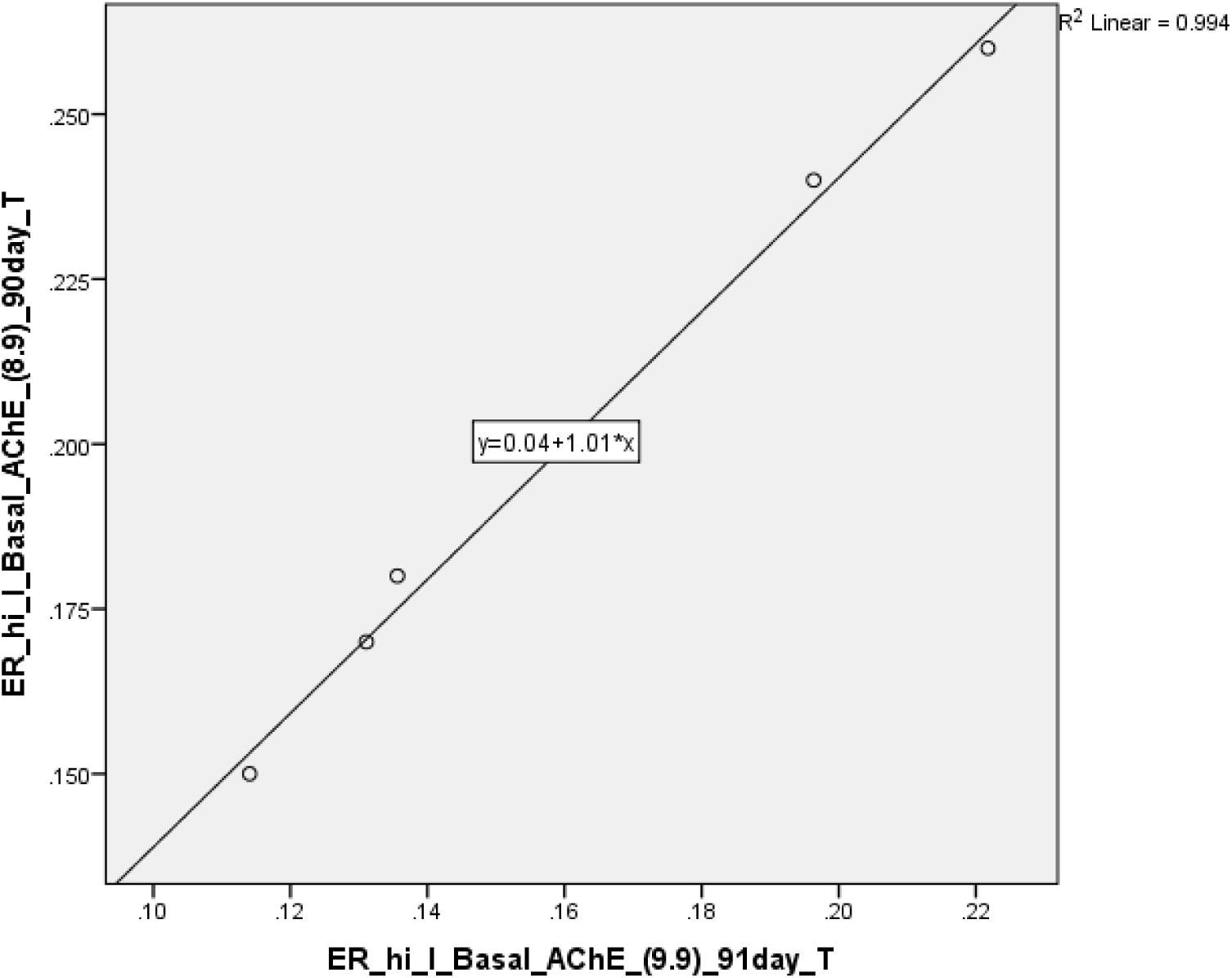
Linear stepwise regression correlation for Basal AChE activity at 0 and 91 day of freezing, in ER samples of Test group (R2=0.994) confirmed with cross validation methodology for 80%samples (p<0.01, R=0.997**)

**Graph 4.7.**
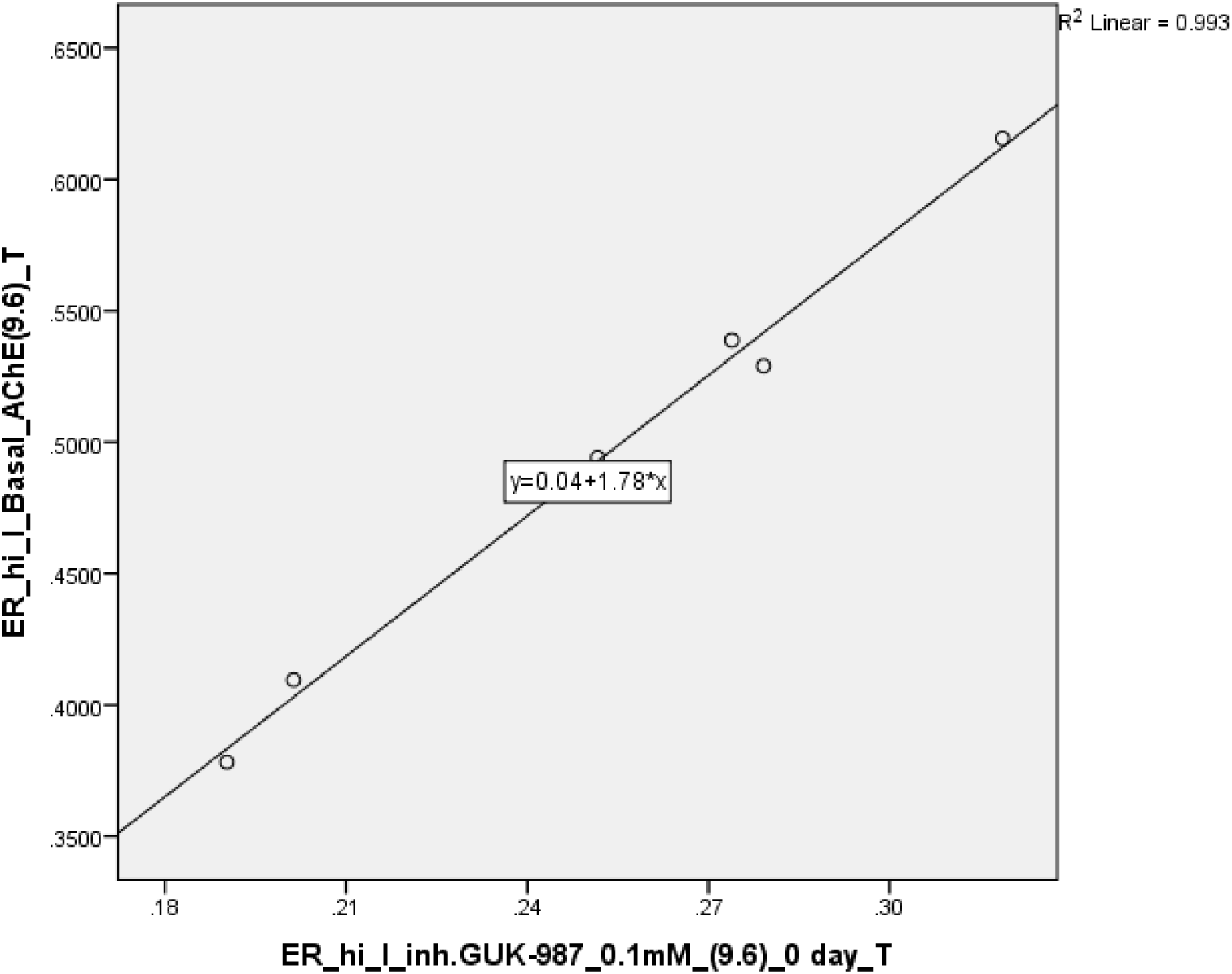
Linear stepwise regression correlation for Basal AChE activity and inhibitor GUK 987 efficacy at 0 day of freezing, in ER samples of Test group (R2=0.993) confirmed with cross validation methodology for 80%samples (p<0.01, R=0.996**)

**Graph 4.8.**
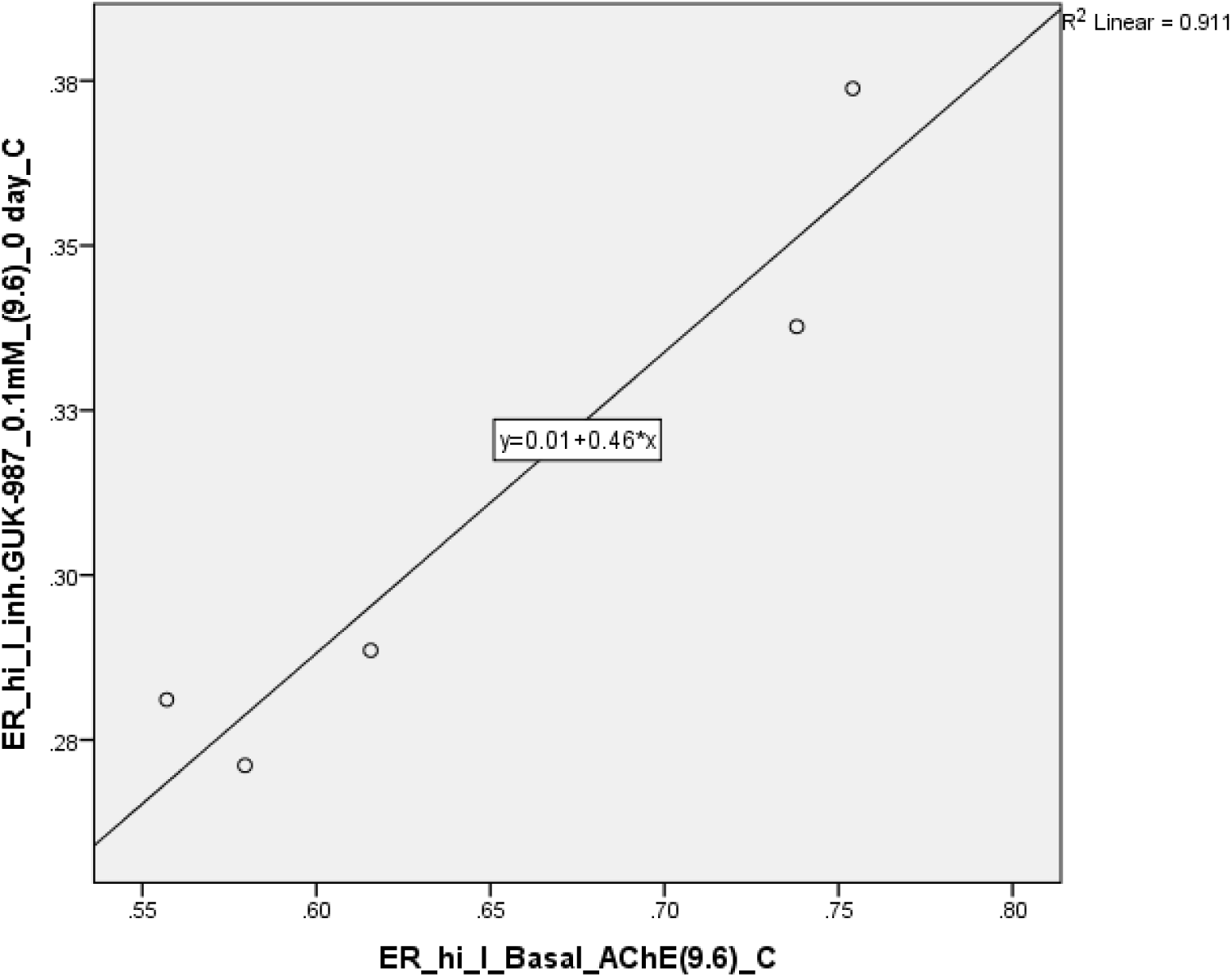
Linear stepwise regression correlation for Basal AChE activity and inhibitor GUK 987 efficacy at 0 day of freezing, in ER samples of Control group (R2=0.911) confirmed with cross validation methodology for 80%samples (p<0.01, R=1.000**)

**Graph 4.9.**
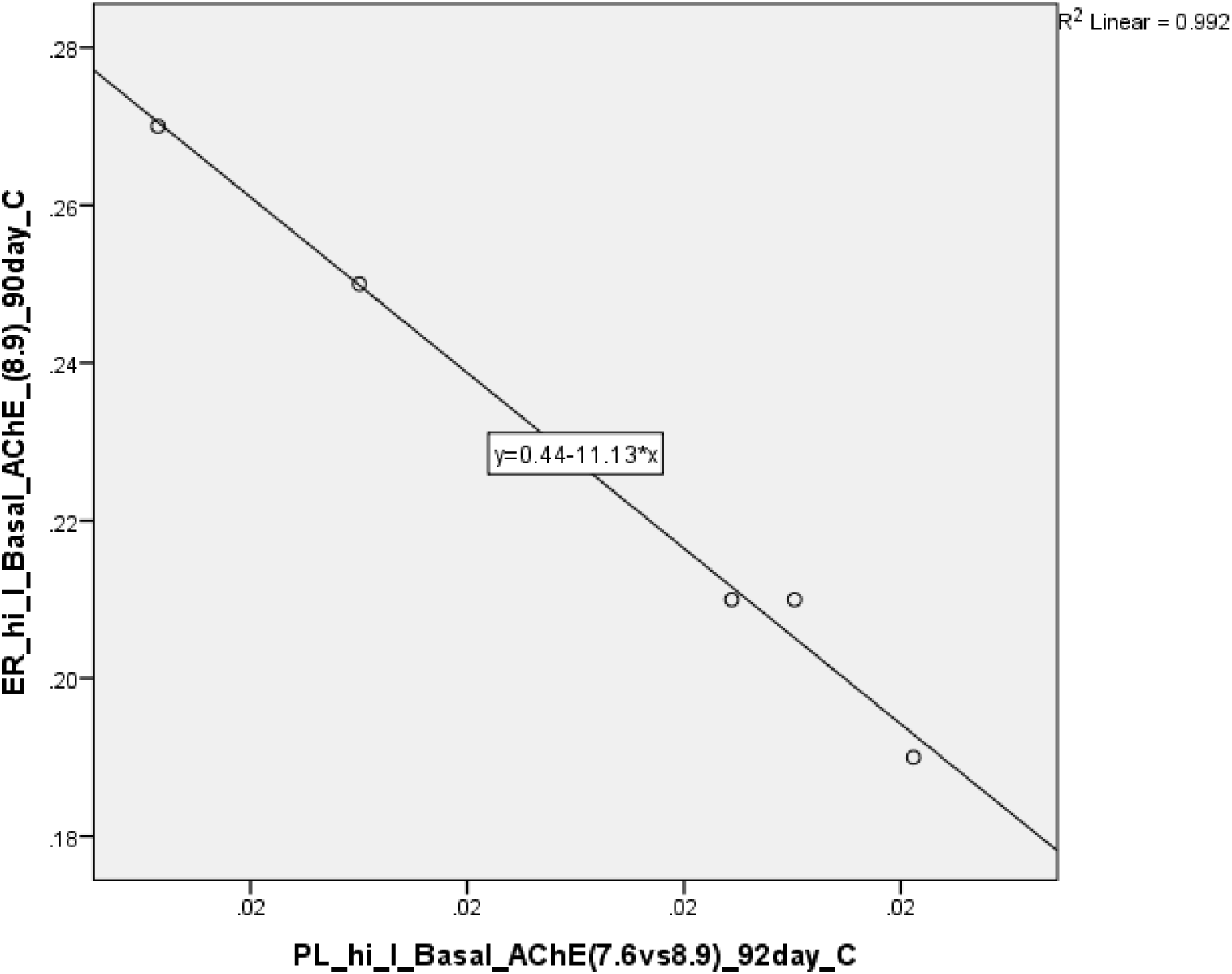
Linear stepwise regression correlation for Basal AChE activity at 90 and 92 day of freezing, in ER and PL samples of Control group (R2=0.992), confirmed with cross validation methodology for 80%samples (p<0.01, R=0.970**)

**Graph 4.10.**
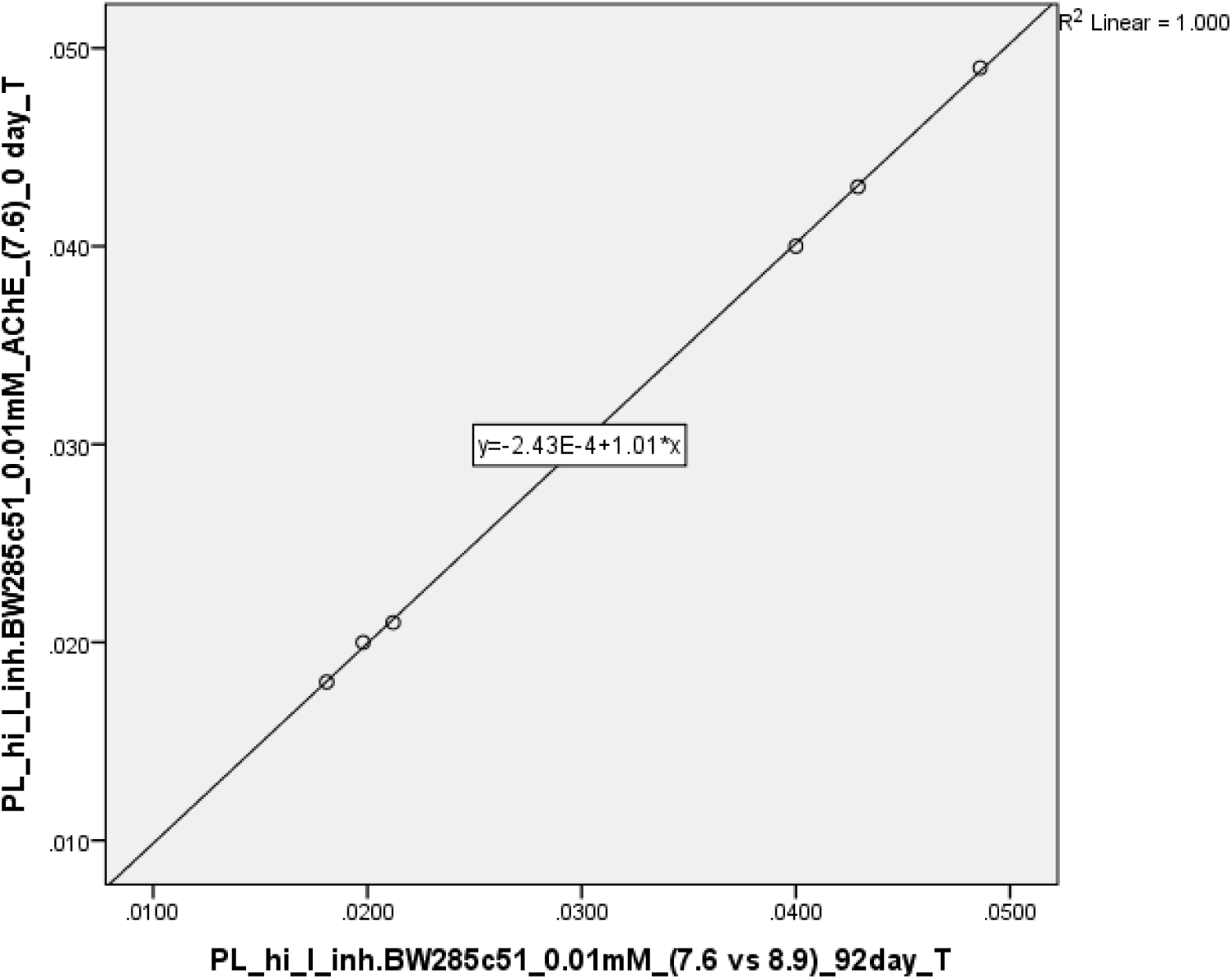
Linear stepwise regression correlation for inhibitor BW284c51 efficacy at 0 and 92^nd^ day of freezing in PL samples of Test group (R2=1.000), confirmed with cross validation methodology for 80%samples (p<0.01, R=1.000**)

**Graph 4.11.**
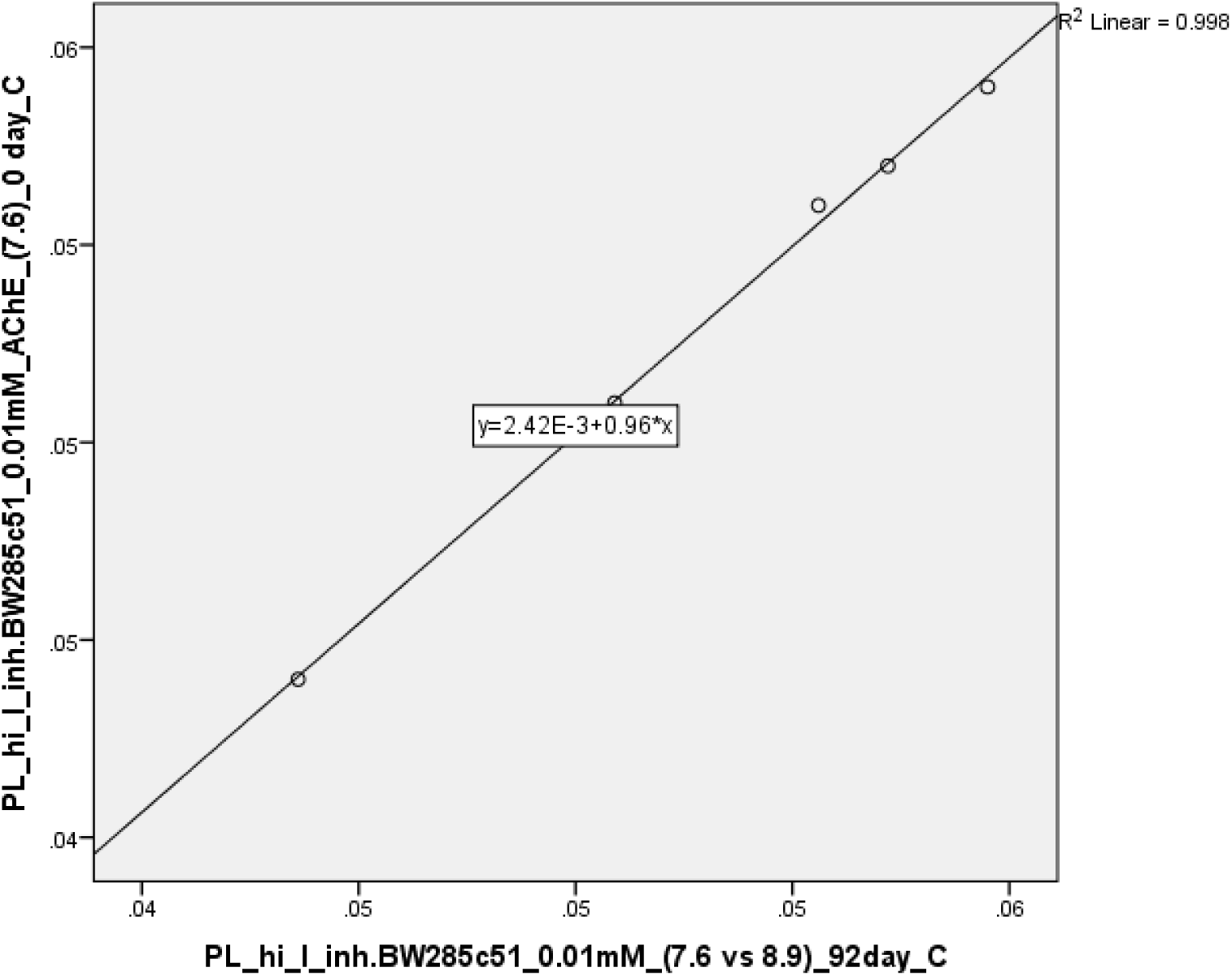
Linear stepwise regression correlation for inhibitor BW284c51 efficacy at 0 and 92^nd^ day of freezing in PL samples of Control group (R2=0.998), confirmed with cross validation methodology for 80% samples (p<0.01, R=1.000**)

**Graph 4.12.**
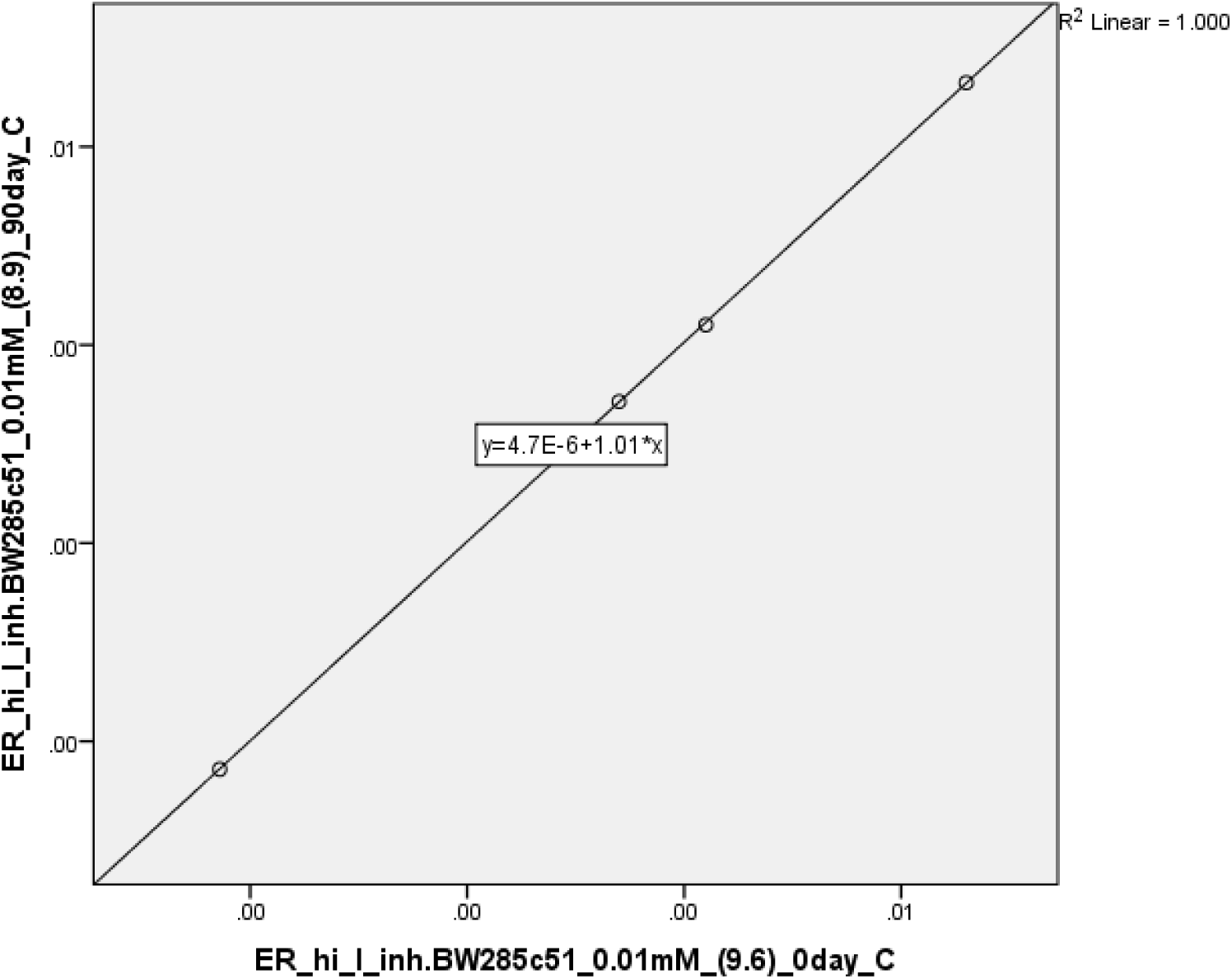
Linear stepwise regression correlation for inhibitor BW284c51 efficacy at 90 and 0 day of freezing in ER samples of Control group (R2=1.000), confirmed with cross validation methodology for 80%samples (p<0.01, R=1.000**)

In order to see if there is **non-linear relationship** among our variables, study also included **enter and binary logistic regression analysis**.

##### 4.2.3.2 Enter regression and binary logistic regression

When **gender or participant** group are set as a dependent variable, there are no results with statistical significant correlation among variables in blood samples. Results from enter and binary logistic regression analysis indicate absence of non-linear relationship among variables.

#### 4.2.4 Paired T test

**Two tailed, paired sample T test** was used to assess the presence of statistical significant correlation between variables for **water intake and storage time** with 95% confidence interval. Results from **Paired T test**, revealed statistical significant correlation (p<0.01) between the next pairs:

***Protein concentration* in** Test group, **PL samples** for day 0 vs 92, 0 vs 93; **ER sample** for day 0 vs 91; PL vs ER for day 0, 92vs 90, 93 vs 91, 93 vs 91. In <u>control</u> group, PL samples for day 0 vs 92, 0 vs 93, **ER samples** for day 0 vs 90; PL vs ER samples for day 0, 92vs90, 93 vs 91.

- ***Basal enzyme AChE activity* in** Test group, **PL samples** for day 0 vs 92; **ER sample** for day 0 vs 90; 0 vs 91, 90 vs 91. Moreover, in **PL samples** correlation is presented for *basalAChE activity vs inhib*.*BW* for day 0, 92; *basal AChE activity and inh. GUK 987* for 92 vs 93 day; *inhibitorBW284c51 vs inh*.*GUK-987* for 92 vs 93 day. **In ER samples** correlation is presented for basal AChE activity vs inh.BW284c51 for day 0, 90; basal AChE activity and inhib.GUK 987 for day 0, 91. Basal enzyme AChE activity in Control group is seen for PL samples during days 0 vs 92, ER samples during 0 vs 90 day. PL vs ER samples during day 0 vs 0; 92 vs 91
- ***Inhibitor efficasy BW284c51* in** Test group, **PL vs ER samples** for day 0, 92 vs 90. In **control group** Inhibitor BW vs Inhib GUK in PL samples showed correlation between 92 vs 93^rd^ day. PL vs ER samples during day 92 vs 90.
- ***Inhibitor efficasy GUK-987* in** Test group, **ER sample** for day 0 vs 91;PL vs ER samples for 93 vs 91 day. Inhibitor efficacy GUK 987 **in Control group**, ER samples for 0 vs 91 day; PL vs ER samples for 93 vs 91 day.

#### 4.2.5 One way ANOVA

F statistics, showed presence of statistical significant difference (p<0.01) for **protein concentration in ER samples during 90**^**th**^ **day**, when comparison is made between test and control group. Differences in the results between Paired T test and One way ANOVA, can be explained by usage of different formulas for deriving error estimates

### 4.3 Building inhibitor calibration curve

**Inhibitor GUK-987** is predominantly BChE inhibitor, but based on the above result it is possible to unspecific inhibit AChE enzyme activity. Furthermore, since **inhibitor GUK-987** is more efficient in PL samples, this study checks concentration gradient range and determines the most efficient concentration for inhibition of enzyme AChE activity after sample storage. These results are logical, since there is more BChE enzyme in PL samples. However, we put substrate for enzyme AChE in order to see if it is possible to inhibit unspecific AChE activity. Moreover, this study included additional BChE inhibitor, **inhibitor GDK-510**, with the same research aim. Literature regarding two inhibitors lack, so based on our current research knowledge this is the first study conducting this type of research and presenting results to the scientific community. **Method development and building calibration curve** for determining the most efficient concentration of unspecific enzyme AChE inhibition is assessed. All sampling was done 29.06 and inhibitor efficacy is assessed after 6, 8 and 9 weeks of storage time at −20°C in the freezer.

Workflow timeline presented in ***Chart 1***, shows development of methodology and understanding inhibition efficacy of enzyme AChE in PL samples with **GUK 987 and GDK 510** inhibitors.

**Chart 1.**
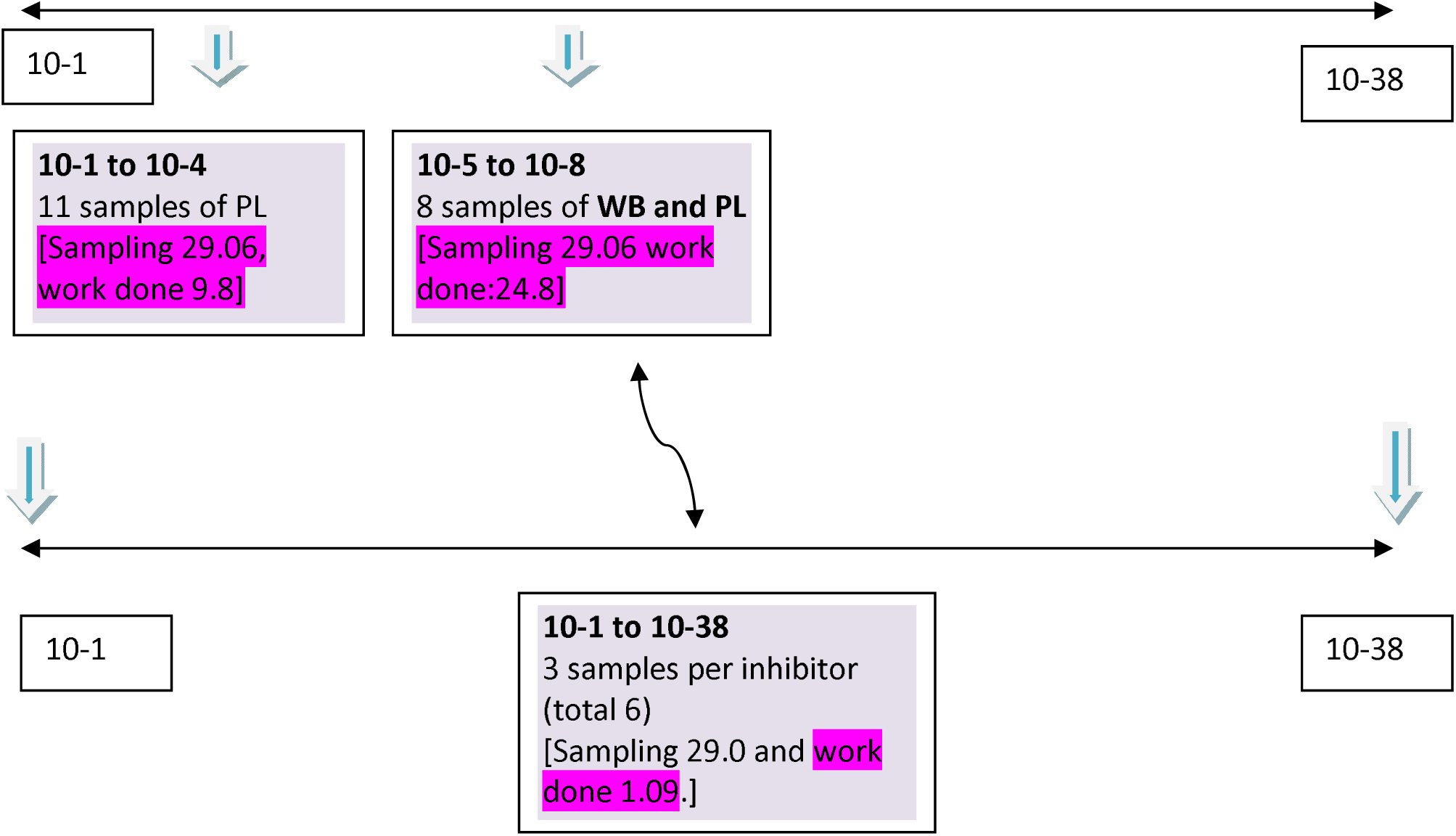
Timeline of workflow. Building inhibitor GUK 987/Gdk-510 calibration curve (method development).

In order to check the inhibition efficacy of inhibitor in PL and WB samples, we performed the analysis on diverse concentration ranges. At first we analyzed all 11 PL samples for the range of concentration (10-1 to 10-4). Secondly, we analyzed 8 WB and PL samples for the range of concentrations (10-5 to 10-8). Inhibitor **GUK 987 and GDK-510** inhibits AChE activity in **PL and WB** samples. Inhibitor showed the most efficient inhibition in **PL samples**. Based on this notion, calibration curve efficacy of inhibitor **GUK-987 and GDK-510** in the broader range **[10-1 to 10-38]** was determined for **6 samples (3 samples per 1 inhibitor)** after 9 weeks of storage. Minimal number of samples was used. Basal enzyme AChE activity was measured using AChE substrate. However, inhibitor GUK 987 and GDK −510 are inhibitors of BChE enzyme. So we measured unspecific inhibition of **enzyme AChE** with BChE inhibitors. In order to see if the effect will cause inhibition of enzyme activity in plasma samples, calibration curve was done on **total 6 plasma samples** [PL_Test samples=GUK 987 and PL_Control samples=GDK 510]. Concentration gradient [10-1 to 10-38] is made from **1mM inhibitor stock**. Since we measured unspecific inhibition of enzyme AChE in PL samples, results indicate that inhibitor GUK-987 is more efficient in Test group samples, where water intake was higher before sampling while inhibitor GDK-510 is more efficient in control group samples, where water intake was less before sampling.

Results show that the most efficient inhibition concentration in Plasma samples for inhibitor GDK-987 is **10-5mM** while for inhibitor GDK-510 is **10-3mM**. Calibration curve with concentration logarithm is presented in the Graph 5.

**Graph 5.**
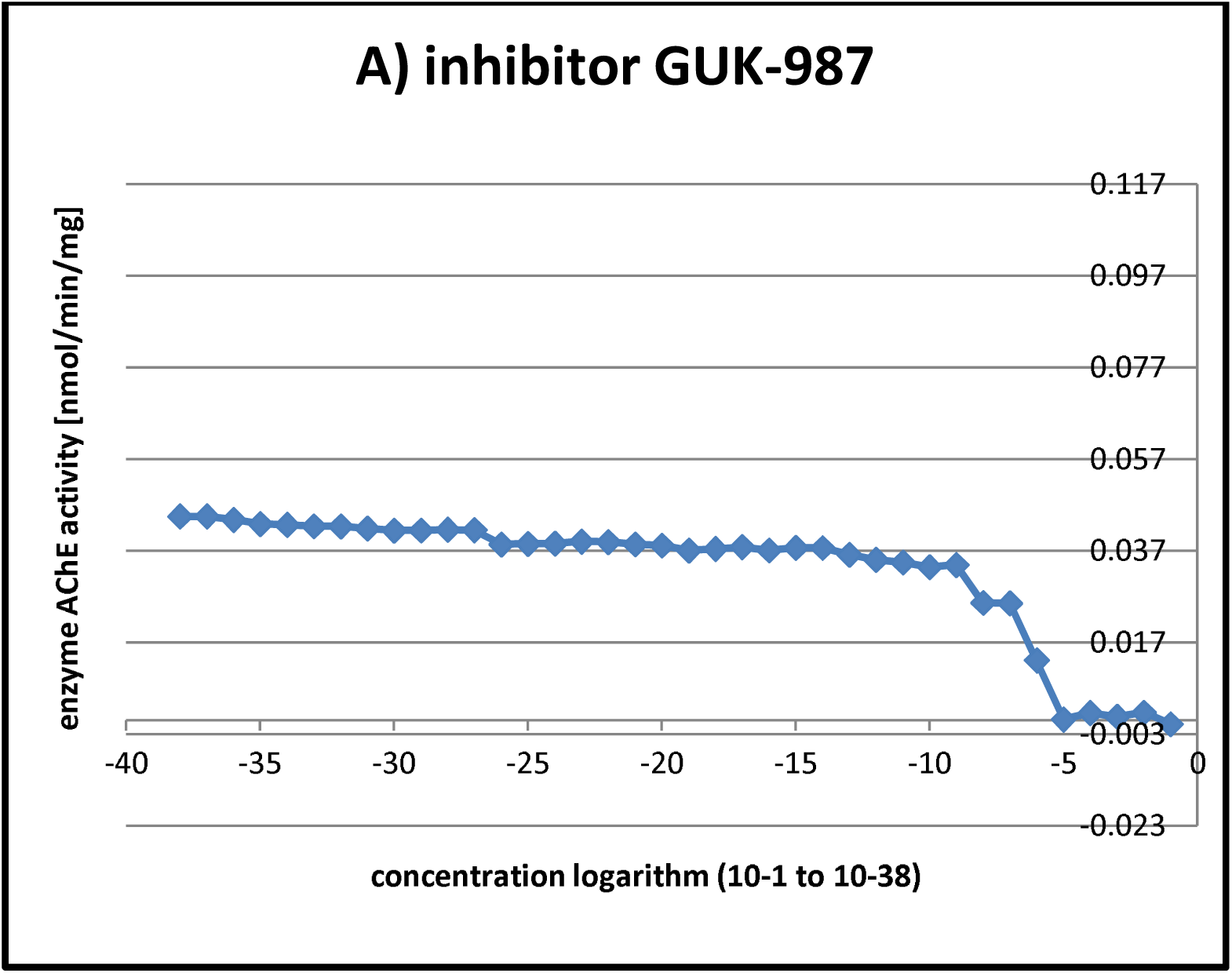
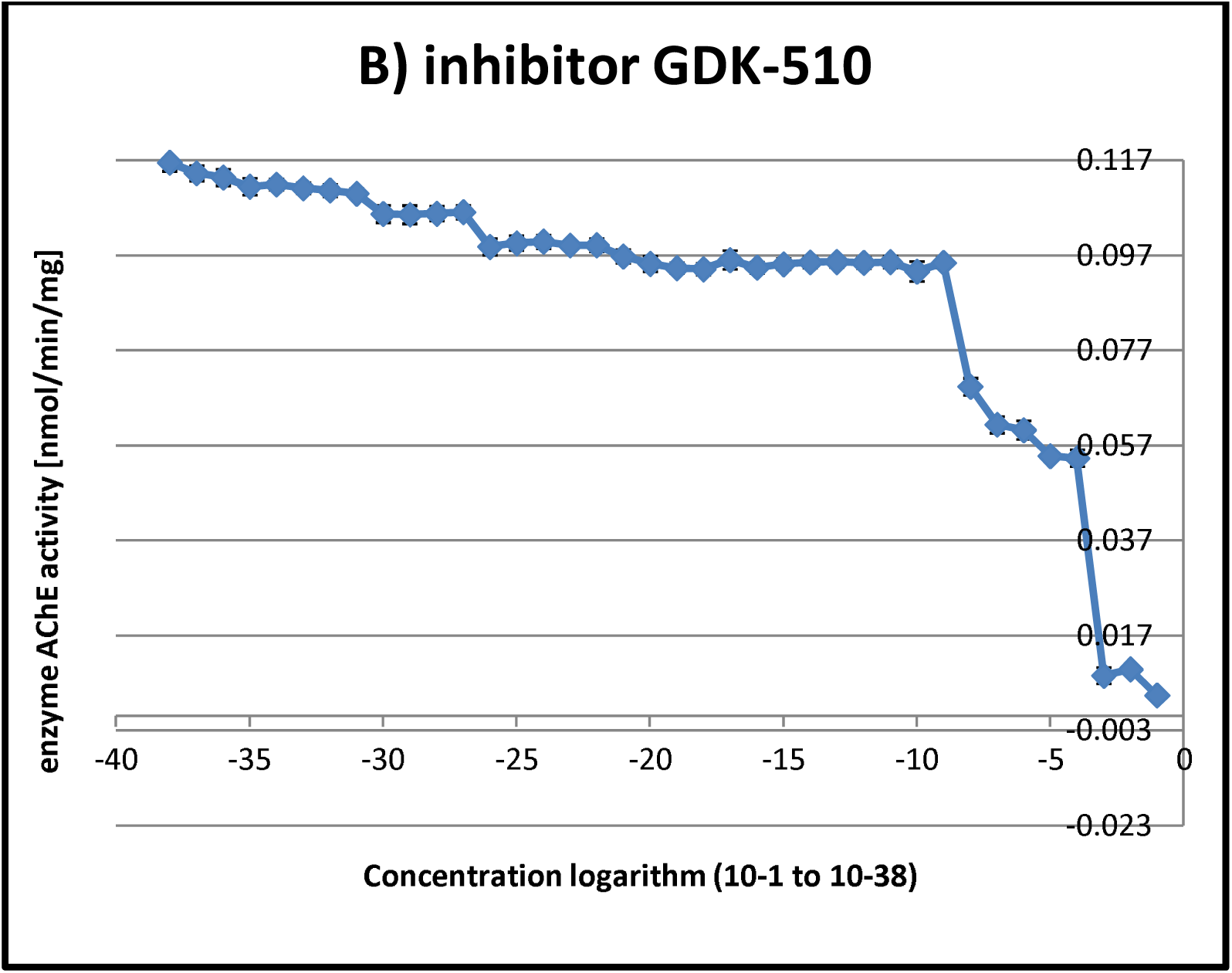
Inhibitor efficacy of inhibition in plasma samples expressed in logarithm gradient concentration. A) Inhibitor GUK-987; B) Inhibitor GDK-510.

## 5. Discussion

Sample management is inevitable practice for clinical and non clinical studies [21]. Understanding sampling procedure from collection, handling, storage, shipment, pre analysis and post analysis storage and disposal is of key importance for proper result interpretation [21]. Type of used anticoagulants, volume, light sensitivity, containers and labeling gives basic information to the researcher for future analysis [22]. Sometimes, blood samples are frozen and may be transported to longer distance without any consequences [23]. Prolonged storage from few months or years is possible for blood specimens before processing, depending on the purpose of analysis [23]. Inappropriate long term storage can affect negatively on sample quality and results. Effect of prolonged storage in blood samples on different temperatures has been done [24]. There have been debates regarding stability of samples at lower and higher temperature [23].

Moreover, drug therapy has enormous cost on the market per year, and because of that long term storage is important for maintaining sample integrity and properties [25]. Control and access to samples provide security and reliability of service quality [23]. Enable long term analytical testing, record of the analysis and pharmacovigilance report [21, 23] Sample storage time can have an effect on sample **quality and final result** interpretation [21]. Depending on the analyzed analites results vary. Because of that it is **important and challenging** to get a grasp of the AChE enzyme activity change in human blood in healthy individuals depending on water intake and storage time. All conditions should be stable and quality control samples should be used to minimize false results, taking into the account diverse influencing factors [26]. Factors influencing biomarker composition are days of freezing, season and month of the year for sample collection, participant age and gender, number of participants, sample type, freezer and thaw cycles, intraindividual factors and environmental factors (organophosphate poisoning, sun-light and ultra violet radiation) [27]. All this factors influence enzyme AChE activity in blood samples except age and gender [28-34]. Stability of biomolecules is important for short term (quantity of molecules) and long term stability (collection of samples for rare diseases, monitoring individual variance in longitudinal studies over extended time periods, and for retrospective studies with samples stored over different time-spans or storage methods) [27].

Establishing evidence of enzyme change can help researchers in accommodating the effect of a therapy based on the results. The proposed methodology is cost effective, simple, fast and practical for routine analysis. When assessing efficacy of new drug targets it is important to take into account storage time on analyzed blood parameters among other potential influencers. Usage of diverse statistical test made in the results section, confirms the need for statistical methodology refinement. Depending on the statistical tests and usage of different formulas for deriving error estimates, this study indicates that **protein concentration and enzyme AChE activity** decreases in PL and ER samples with statistical significance during storage time on −20°C in the freezer. **Inhibitor GUK 987 (0.1mM)** is more efficient in plasma and erythrocyte samples with higher water intake, while **inhibitor BW284c51 (0.01mM)** is more efficient in Erythrocyte samples. Moreover, this study compared the effect of 92 days of storage at −20°C in the freezer utilizing **Acetylthiocholine chloride** as substrate, meaning it measured AChE activity in PL samples as well. Other studies analyzed AChE changes in activity of a **few hours to few days till one month**, indicating also statistical significant decrease in the enzyme activity rate in human blood samples [35, 36].

Discrepancy of the enzyme AChE stability exists among literature. Stability of enzyme activity is held for 7, 14 and 31 days unchanged in ER samples, with diverse decrease intensity after proposed storage freezing, depending on used methodology [16, 17 37, 38]. Human serum samples, showed absence of AChE activity change after one year of storage [18]. Based on other study, difference in the enzyme AChE activity between 34^st^ and 50^th^ day of storage in ER samples are the same, reaching 10% decrease [35, 37]. Results from this study indicate higher decrease of basal enzyme AChE activity in Control group, to 28.57% and 25% activity in Female/Males after 92 days of storage in PL samples and 25.57% /33.33% in ER samples of Male participants in Test and Control group. We can conclude that depending on the water intake there is higher decrease of AChE activity in PL samples in comparing to ER samples.

Usually, researchers use PL samples to measure BChE activity, utilizing butyrlcholine iodide as substrate. Results measured no change in enzyme AChE activity after 14 months of storage [19]. This study used substrate for enzyme AChE and measured unspecific inhibition with BChE inhibitors GUK-987 and GDK-510 on enzyme AChE activity. Results suggest unspecific influence of BChE inhibitors on enzyme AChE activity. Other studies utilized animal model systems, like meat from animals used as food, horses and dogs, for AChE activity change after proposed day of freezing samples at −20°C in the freezer. Gain results are similar to the above, although they used human samples [13-15]. Based on the above notion, available literature lack and indicate absence of practical information regarding **water intake prior sampling and effect of freezing time** on protein concentration, basal AChE activity and inhibitor efficacy in PL and ER samples. This study continues the previous study S. Jovicic regarding the effect of water intake on enzyme activity. Moreover, results made a new contribution regarding calibration of inhibitor efficacy and how adequate protocols can be made.

Results from this study are in the accordance of .mention studies, regarding significant change in AChE activity after 92 days of storage at −20°C. However, the percentage of decrease could not be followed from their results. In our study the fall of protein concentration, basal AChE activity and inhibitor efficacy is significant and it differs based on sample type. After 92 days of storage, Plasma samples showed highest decrease of **protein concentration** in F test group (48%) and M control group (40.85%), **basal AChE activity** in M/F Test group (40%/33.3%). After 90 days of storage, Erythrocyte samples showed highest decrease of protein concentration and basal AChE activity in M/F group (80%/85.6% of protein concentration and 40.81%/33.33% in basal AChE activity). Inhibition efficacy of 100% for all genders and sample groups is seen in all genders and sample groups for Plasma samples and inhibitor GUK-987, 0.1mM and ER samples and inhibitor BW284c51, 0.01mM for ER samples. The methodology Elman is shown to be sensitive and detect significant decrease in protein concentration, enzyme activity and inhibitor efficacy depending on the water intake before blood samples were collected.

## 6. Conclusion

Temperature of freezing and days of storage are important strategy for proper sample analysis and reuse of available material of the participants. Water intake before sampling can affect the results of potential biomarker depending on the days of sample storage.

This is the first study investigating the effect of prolonged freezing of maximum 92 days on protein concentration, enzyme AChE activity in ER and PL samples, by using diverse statistical methodology for deriving conclusion in healthy participants with diverse water intake before sampling. Main finding from this study was that water intake before sampling of biological material can have statistical significant influence on protein concentration, enzyme AChE activity and inhibitor efficacy in Plasma and Erythrocyte blood samples. Methodology is sensitive and cost effective for further clinical studies. **Disadvantage** of the study is small number of samples and gender discrepancy. However, this disadvantage is minor, since other studies published similar data on smaller participant number [16]. **Advantage** of this study is estimating the prior effect of water intake before sampling on protein concentration, enzyme AChE activity and inhibitor efficacy decline during maximum 92 days storage time at −20°C in the freezer and assessing applicability of the results for additional clinical studies.

Future work should take into the account higher number of samples, participants during diverse physiological states in health and disease in order to better assess the influence on establishing range activity decline of investigated parameters.

## Data Availability

Data available from Table 1

## List of abbreviations

A AChE: Acetilcholine esterase, (sinonim: blood, erythrocyte cholinesterase)
B BChE: Butyrylcholinesterase (sinonim: plasmatic, pseudocholinesterase)
C ChE: Cholinesterase enzymes
C: Control
E ER: Erythrocites
F FDA: Food Drug Agency
L LE: Leucocyte
P PL: Plasma
Pt: Platelet
T T: Test
W WB: Whole blood

## Declaration

### Ethical approval

Experiments were performed according to **ethical standards** and with written consent of the blood donors. The study was given ethical permission from the National Medical Ethics Committee (number 82/07/14). Experimental part of the experiment was done at the Biotechnical Faculty, University of Ljubljana, Slovenia, within the project **J5-7098** and **CEEPUS free mover PhD student program mobility** with the University of Belgrade, Serbia.

### Consent for publication

I, Snežana Jovičić (PhD student, Faculty of Biology, University of Belgrade, Serbia), as an only author of this systematic review, give my consent for contained information regarding this manuscript and information about myself to be published in this Journal.

### Availability of data material

Data available from Table 1

### Authors contribution

Author (Snežana Jovičić, PhD student at the Faculty of Biology, University of Belgrade, Serbia), originated idea for this study along with paper and data collection, analysis, interpretation and writing. Snežana Jovičić made all the effort and contribution by herself for the present work and ensures that questions related to the accuracy or integrity of any part of the work are appropriately investigated, resolved, and documented. She is solely responsible for the quality of this produced work.

Author (Snežana Jovičić) approves the final version of the presented manuscript for submission.

Snežana Jovičić, **confirms** that this work is original and has not been published elsewhere, nor is it currently under consideration for publication elsewhere.

#### Aknowledgment

I want to acknowledge the kind support of my CEEPUS freemover mobility mentor from Slovenia, prof. dr. **Damjana Drobne**, who gave me the opportunity to work on this subject by ensuring me samples and place between research roof top. She was kind to accept me to work with her on a project, to finalize my PhD thesis through CEEPUS freemover mobility network, and establish close collaboration between Universities. She introduced me closer to academia and scientific research. Thanks to her, I was taught and I learn scientific independence. I also want to acknowledge my dear **colleges** from the **Biotechnical** faculty, M.Sc Alenka Malovrh, PhD student Neža Repar, and other team members of Bionanoteam who introduced me to practical scientific work in laboratory and student mentoring. **Prof. dr. Veronika Kralj Iglic**, PI, who gave me information about the project J5-7098 "Assessment of blood parameters and extracellular vesicles for optimisation of sport results” on which I worked through CEEPUS freemover mobility.

Moreover, I want to thank all the people who contributed to my education, people being a part of my life during some period and my parents for the continuous given support and love. Thank everyone who ever came across my road of life. Just to let you know, earth is round and I will never change. I am sending you a lot of hugs and kisses. Thank you all for being a part of this lovely Snežana (Snow White) story, my life story. Life writes novels.

## Author’s information

**Snežana Jovičić**, a *PhD student* is born 17.10.1990 in Belgrade, Serbia. She finished two high schools, Medical and Musical high school. Academic qualifications: enrolment of PhD studies, Genetics (2014). Finished **M.Sc. Human Molecular biology, (2014) and B.Sc. Molecular Biology and Physiology (2013)**. Snežana is hardworking, creative, communicative and charismatic person, capable of working in diverse situations, effectively managing time. Adore solving complex intellectual problems and communication with diverse personalities. During spare time, loves activities such as swimming, rollerblades, playing violoncello, singing, classical music, climbing, dancing salsa and Latin dances. Moreover, spending free quality time with family and friends. Very important part of everyday life is work on personal and professional development. Reading books in psychology, archeology, art, travel and meeting new people, cultures and ways of living. Areas of scientific interest are connected to human diseases (Oncology, Inflammation and Cardiovascular Diseases), activation of molecular pathways, gene expression, clinical studies, farmacovigilance, connecting medicine and science, academic reading, writing, data analysis. Personal interest is connected in increasing quality of human healthcare and creating widely applicable knowledge.

## Reference

1. National Research Council (US) Committee on Prudent Practices in the Laboratory (2011). Prudent Practices in the Laboratory: Handling and Management of Chemical Hazards: Updated Version. Washington (DC): National Academies Press (US); Laboratory Security. Available from: https://www.ncbi.nlm.nih.gov/books/NBK55881/ [Accessed:29.09.2020]

2. Vaught JB, Henderson MK. Biological sample collection, processing, storage and information management. IARC Sci Publ. 2011; 163:23-42. PMID: 22997855

3. World Health Organization (2011). Laboratory Quality Management System. Handbook. Available at: https://www.who.int/ihr/publications/lqms_en.pdf [Accessed: 29.09.2020]

4. Hammerling JA. A Review of Medical Errors in Laboratory Diagnostics and Where We Are Today. Laboratory Medicine. 2012; 43 (2): 41–44. https://doi.org/10.1309/LM6ER9WJR1IHQAUY

5. Barr AJ. The biochemical basis of disease. Essays Biochem. 2018; 62 (5): 619–42. https://doi.org/10.1042/EBC20170054

6. Jackson M, Marks L, May GHW, Wilson JB. The genetic basis of disease. Essays Biochem. 2018; 62(5):643–723. https://doi.org/10.1042/EBC20170053

7. Lima-Oliveira G, Lippi G, Salvagno GL, Picheth G, Guidi GC. Laboratory Diagnostics and Quality of Blood Collection. J Med Biochem. 2015; 34(3):288–94. https://doi.org/10.2478/jomb-2014-0043

8. do Amaral RJ, da Silva NP, Haddad NF, Lopes LS, Ferreira FD, Filho RB, Cappelletti PA, de Mello W, Cordeiro-Spinetti E, Balduino A. Platelet-Rich Plasma Obtained with Different Anticoagulants and Their Effect on Platelet Numbers and Mesenchymal Stromal Cells Behavior In Vitro. Stem Cells Int. 2016; 2016:7414036. http://doi.org/10.1155/2016/7414036

9. Dvir H, Silman I, Harel M, Rosenberry TL, Sussman JL. Acetylcholinesterase: From 3D Structure to Function. Chemico-Biological Interaction. 2010; 187 (1-3):10–22. http://dx.doi.org/10.1016/j.cbi.2010.01.042

10. Naik RS, Doctor BP, Saxena A. Comparison of methods used for the determination of cholinesterase activity in whole blood. Chem Biol Interact. 2008; 175(1-3):298–302. http://doi.org/10.1016/j.cbi.2008.05.002

11. Durrant AR, Tamayev L, Anglister L. Serum cholinesterases are differentially regulated in normal and dystrophin-deficient mutant mice. Front Mol Neurosci. 2012; 5:73. http://doi.org/10.3389/fnmol.2012.00073

12. Murray AP, Faraoni MB, Castro MJ, Alza NP, Cavallaro V. Natural AChE Inhibitors from Plants and their Contribution to Alzheimer’s Disease Therapy. Curr Neuropharmacol. 2013; 11(4):388–413. http://doi.org/10.2174/1570159X11311040004

13. Plumlee KH, Richardson ER, Gardner IA, Galey FD. Effect of time and storage temperature on cholinesterase activity in blood from normal and organophosphorus insecticide-treated horses. J Vet Diagn Invest. 1994; 6(2):247–9. http://doi.org/10.1177/104063879400600217

14. Tecles F, Gutiérrez PC, Martínez SS, Cerón JJ. Effects of different variables on whole blood cholinesterase analysis in dogs. J Vet Diagn Invest. 2002; 14(2):132–9. http://doi.org/10.1177/104063870201400207

15. Askar KA, Kudi AC, Moody AJ. Comparison of two storage methods for the analysis of cholinesterase activities in food animals. Enzyme Res. 2011; 2010:904249. http://doi.org/10.4061/2010/904249

16. Oliveira-Silva JJ, Alves SR, Inacio AF, Meyer A, Sarcinelli PN, Mattos RC, Ferreira MF, Cunha JC, Moreira JC. Cholinesterase activities determination in frozen blood samples: an improvement to the occupational monitoring in developing countries. Hum Exp Toxicol. 2000; 19(3):173–7. http://doi.org/10.1191/096032700678827762

17. Gupta VK, Pal R, Siddiqi NJ, Sharma B. Acetylcholinesterase from Human Erythrocytes as a Surrogate Biomarker of Lead Induced Neurotoxicity. Enzyme Res. 2015; 2015:370705. http://doi.org/10.1155/2015/370705

18. Huizenga JR, van der Belt K, Gips CH. The effect of storage at different temperatures on cholinesterase activity in human serum. J Clin Chem Clin Biochem. 1985; 23(5):283–5. http://doi,org/10.1515/cclm.1985.23.5.283

19. Turner JM, Hall RA, Whittaker M, Kricka LJ. Effects of storage and repeated freezing and thawing on plasma cholinesterase activity. Ann Clin Biochem. 1984; 363–5. http://doi.org/10.1177/000456328402100504

20. Henchman RH, Tai K, Shen T, McCammon JA. Properties of water molecules in the active site gorge of acetylcholinesterase from computer simulation. Biophys J. 2002; 82(5):2671–82. http://doi.org/10.1016/S0006-3495(02)75609-9

21. Redrup MJ, Igarashi H, Schaefgen J, Lin J, Geisler L, Ben M’Barek M, Ramachandran S, Cardoso T, Hillewaert V. Sample Management: Recommendation for Best Practices and Harmonization from the Global Bioanalysis Consortium Harmonization Team. AAPS J. 2016; 18(2):290–3. http://doi.org/10.1208/s12248-016-9869-2

22. Tuck MK, Chan DW, Chia D, Godwin AK, Grizzle WE, Krueger KE, Rom W, Sanda M, Sorbara L, Stass S, Wang W, Brenner DE. Standard operating procedures for serum and plasma collection: early detection research network consensus statement standard operating procedure integration working group. J Proteome Res. 2009; 8(1):113–7. http://doi.org/10.1021/pr800545q

23. van de Merbel N, Savoie N, Yadav M, Ohtsu Y, White J, Riccio MF, Dong K, de Vries R, Diancin J. Stability: recommendation for best practices and harmonization from the Global Bioanalysis Consortium Harmonization Team. AAPS J. 2014; 16(3):392–9. http://doi.org/10.1208/s12248-014-9573-z

24. Bulla A, De Witt B, Ammerlaan W, Betsou F, Lescuyer P. Blood DNA Yield but Not Integrity or Methylation Is Impacted After Long-Term Storage. Biopreserv Biobank. 2016; 14(1):29–38. http://doi.org/10.1089/bio.2015.0045

25. Pagès A, Foulon S, Zou Z, Lacroix L, Lemare F, de Baère T, Massard C, Soria JC, Bonastre J. The cost of molecular-guided therapy in oncology: a prospective cost study alongside the MOSCATO trial. Genet Med. 2017; 19(6):683–90. http://doi.org/10.1038/gim.2016.174

26. Burd EM. Validation of laboratory-developed molecular assays for infectious diseases. Clin Microbiol Rev. 2010; 23(3):550–76. http://doi.org/10.1128/CMR.00074-09

27. Enroth S, Hallmans G, Grankvist K, Gyllensten U. Effects of Long-Term Storage Time and Original Sampling Month on Biobank Plasma Protein Concentrations. EBioMedicine. 2016; 12:309–14. http://doi.org/10.1016/j.ebiom.2016.08.038

28. Oliveira-Silva JJ, Alves SR, Inacio AF, Meyer A, Sarcinelli PN, Mattos RC, Ferreira MF, Cunha JC, Moreira JC. Cholinesterase activities determination in frozen blood samples: an improvement to the occupational monitoring in developing countries. Hum Exp Toxicol. 2000; 19(3):173–7. http://doi.org/10.1191/096032700678827762

29. Altinyazar V, Sirin FB, Sutcu R, Eren I, Omurlu IK. The Red Blood Cell Acetylcholinesterase Levels of Depressive Patients with Suicidal Behavior in an Agricultural Area. Indian J Clin Biochem. 2016; 31(4):473–9. http://doi.org/10.1007/s12291-016-0558-9

30. Jennings NA, Pezzementi L, Lawrence AL, Watts SA. Acetylcholinesterase in the sea urchin Lytechinus variegatus: characterization and developmental expression in larvae. Comp Biochem Physiol B Biochem Mol Biol. 2008; 149(3):401–9. http://doi.org/10.1016/j.cbpb.2007.10.014

31. Zlatkovic M, Krstic N, Boskovic B, Vucinic S. Determination of reference values of acetyl and Butyryl cholinesterase activities in Serbian healthy population. Military Medical Journal. 2016; 74(00):101–101. http://doi.org/10.2298/VSP160303101Z

32. Sánchez Luz Helena, Medina Olga Marcela, Gómez Guillermo, González Clara Isabel Flórez-VargasÓscar. Laboratory genetic-based reference values for cholinesterase activity in a Colombian population: A step forward in personalized diagnostics. Biomédica. 2015; 35 (2): 20–9. http://doi.org/10.7705/biomedica.v35i0.2422

33. Gold J, Shoaib A, Gorthy G, Grossberg GT. The role of vitamin D in cognitive disorders in older adults. 2018. US Neurology. 2018; 14(1): 41–6. http://doi.org/10.17925/USN.2018.14.1.41

34. Rodrigues MV, Gutierres JM, Carvalho F, Lopes TF, Antunes V, da Costa P, Pereira ME, Schetinger MRC, Morsch VM, de Andrade CM. Protection of cholinergic and antioxidant system contributes to the effect of Vitamin D3 ameliorating memory dysfunction in sporadic dementia of Alzheimer’s type. Redox Rep. 2019; 24(1):34–40. http://doi.org/10.1080/13510002.2019.1617514.

35. Crane CR, Sanders DC, Abbott JK (1970). Studies on the storage stability of human blood cholinesterases: I. United states Office of Aviation Medicine. AM 70-4; DOT/FAA/AM-70/4. Available at: https://rosap.ntl.bts.gov/view/dot/20893 [Accessed: 19.09.2020].

36. Freitas Leal JK, Adjobo-Hermans MJW, Brock R, Bosman GJCGM. Acetylcholinesterase provides new insights into red blood cell ageing in vivo and in vitro. Blood Transfus. 2017; 15(3):232–238. http://doi.org/10.2450/2017.0370-16.

37. Worek F, Mast U, Kiderlen D, Diepold C, Eyer P. Improved determination of acetylcholinesterase activity in human whole blood. Clin Chim Acta. 1999; 288(1-2):73–90. http://doi.org/10.1016/s0009-8981(99)00144-8.

38. Muderhwa J, Pfluger K, Olson D, Pee A, Quintana M, Hall S. Study on the Storage Viability of Human Red Blood Cell Cholinesterase. American Journal of Clinical Pathology. 2015; 144:A062. http://doi.org/10.1093/ajcp/144.supl2.062

